# 24-hour sleep-wake regularity and cognitive aging among 74,733 middle-aged and older adults from the US and Europe: The LifeSPAN Consortium

**DOI:** 10.64898/2026.05.22.26353492

**Authors:** Sanne J.W. Hoepel, Alice Albrecht, Jiawen Chen, Lachlan Cribb, Ian M. Danilevicz, Aron S. Buchman, Lisa L. Barnes, David A. Bennett, Suzanne M. Bertisch, Angus Burns, Timothy M. Hughes, Sonia Ancoli-Israel, Andrew Lim, Annemarie I. Luik, Shaun Purcell, Susan Redline, Katie L. Stone, Frank J. Wolters, Qian Xiao, Kristine Yaffe, Stephanie Yiallourou, Meredith L. Wallace, Peng Li, Séverine Sabia, Matthew P. Pase, Yue Leng

**Affiliations:** Department of Psychiatry and Behavioral Sciences, University of California San Francisco, San Francisco, CA, United States; Department of Epidemiology, Erasmus University Medical Centre, Rotterdam, The Netherlands; Turner Institute for Brain and Mental Health, School of Psychological Sciences, Monash University, Melbourne, Australia; School of Public Health and Preventive Medicine, Monash University, Melbourne, VIC, Australia; Université Paris Cité, Inserm U1153, INRAE, Centre for Research in Epidemiology and Statistics (CRESS), EpiAgeing, Paris, France; Rush Alzheimer’s Disease Center, Rush University Medical Center, Chicago, IL, United States; Department of Medicine, Brigham and Women’s Hospital, Boston, MA, United States; Division of Sleep Medicine, Harvard Medical School, Boston, MA, United States; Division of Sleep and Circadian Disorders, Brigham and Women’s Hospital, Boston, MA, United States; Department of Internal Medicine, Wake Forest University School of Medicine, Winston-Salem, NC, United States; Department of Psychiatry, University of California, San Diego, La Jolla, CA, United States; Department of Neurology, University of Toronto, Toronto, Ontario, Canada; Trimbos Institute – The Netherlands Institute of Mental Health and Addiction, Utrecht, The Netherlands; Center for Aging Science, Sutter Health Research Institute, San Francisco, CA, United States; Department of Epidemiology and Biostatistics, University of California, San Francisco, CA, United States; Department of Radiology & Nuclear Medicine, Erasmus University Medical Centre, Rotterdam, The Netherlands; Department of Epidemiology, University of Texas Health Sciences Center at Houston School of Public Health, Houston, TX, United States; Department of Epidemiology & Biostatistics, University of California, San Francisco, CA, United States; Department of Neurology, University of California, San Francisco, CA, United States; Department of Psychiatry, University of Pittsburgh, Pittsburgh, PA, United States; Department of Anesthesiology, Mass General Brigham, Harvard Medical School, Boston, MA, United States

## Abstract

**Importance:** Irregular sleep-wake patterns have been associated with poor health and cognitive outcomes, yet evidence linking 24-hour sleep-wake regularity to cognitive decline or dementia remains inconsistent. Particularly, regularity can be measured as regularity of rest-wake, sleep-wake or overall 24-hour activity, but it is unclear which aspects are most relevant for cognitive aging.

**Objective:** To assess associations of rest-wake, sleep-wake, and 24-hour activity regularity with cognitive decline and dementia risk.

**Design:** Observational prospective study comprised of six US and European cohorts: MrOS (sleep study between 2003-2005, mean follow-up: 7.1 years), Rotterdam Study (2004-2007, 11.6 years), MESA (2010-2013, 8.2 years), MAP (2005-2018, 7.2 years), Whitehall II (2012-2013, 6.9 years), and UKB (2013-2015, 7.9 years).

**Setting:** Cohort-specific estimates were pooled using random-effects meta-analysis. Analyses were done between June 2025 and March 2026.

**Participants:** 74,733 dementia-free adults with multi-day actigraphy were included across cohorts: MrOS (age: 67-96 years, female:0%), MESA (54-95y, female:54.6%), Rotterdam Study (46-98y, female:55.0%), MAP (56-100y, female:77.1%), Whitehall II (59-83y, female:25.9%), and UKB (55-78y, female:55.5%).

**Exposure:** Day-to-day rest-wake regularity (Rest Regularity Index, RRI), day-to-day sleep-wake regularity (Sleep Regularity Index, SRI), and 24-hour activity regularity (Interdaily Stability, IS) were derived from multi-day actigraphy.

**Main Outcome:** Outcomes were risk of dementia and changes in global cognition.

**Results:** Across six cohorts, 1,906 dementia cases occurred among 74,733 participants. After adjusting for demographics, health behaviors, depressive symptoms and cardiovascular comorbidities, each 1-SD higher regularity score was associated with an 9-14% lower dementia risk (pooled hazard ratios: RRI 0.86 95%CI: [0.79-0.95]; SRI 0.87[0.79-0.97]; IS: 0.91[0.88-0.95]). Associations were approximately linear. Age-stratified analyses showed directionally stronger associations among adults aged < 65, although meta-regression did not support an interaction(p > 0.55). Greater regularity was associated with modestly slower decline in global cognition (pooled β per 1-SD higher score of RRI per year: 0.003, 95%CI [0.001-0.006]).

**Conclusions & Relevance:** Greater regularity of rest-wake, sleep-wake, and 24-hour activity rhythms was associated with lower dementia risk and modestly slower global cognitive decline. These findings suggest that 24-hour sleep-wake regularity is a relevant behavioral marker of cognitive aging and may inform future efforts to identify or intervene on early risk.

**Key points:** *Question:* Is regularity of 24-hour sleep-wake activity associated with subsequent risk of dementia and cognitive decline in middle-aged and older adults?

*Findings:* In this individual participant data meta-analysis of 74,733 adults from the US and Europe, those with more regular rest-wake, sleep-wake, and 24-hour activity rhythms had lower risk of dementia and slower decline in global cognition.

*Meaning:* Maintaining regular 24-hour sleep-wake and activity rhythms may be helpful for long-term cognitive health.

## Introduction

Sleep and circadian disturbances are increasingly recognized as prodromal markers and potentially modifiable risk factors for dementia^1,2^. Irregular sleep-wake rhythms have emerged as strong predictors of cardiometabolic disease and mortality^3,4^. Shift work, an extreme model of chronic irregularity, has also been associated with increased dementia risk^5–7^. Irregularity may promote circadian misalignment, thereby disrupting clearance and aggregation of neurodegenerative proteins, immune signaling, metabolic homeostasis, and other processes implicated in dementia pathogenesis^1,8,9^.

Nevertheless, evidence on sleep-wake regularity and cognitive decline or dementia remains inconsistent with studies reporting null^10–14^, positive^15^ (i.e. greater regularity associated with higher risk), and negative^16–18^ (i.e. greater regularity associated with lower risk) associations. One of the reasons for these inconsistencies may be the use of non-equivalent metrics that capture distinct aspects of irregularity^19^. In older adults, among whom daytime sleep or rest is common^20^, sleep-wake regularity is preferentially characterized across the entire 24-hour cycle rather than inferred from nighttime sleep alone. Metrics differ in behavioral domain (sleep-wake patterns, rest-wake patterns, or activity levels) and temporal framing: some capture sequential day-to-day variability (e.g., Sleep or Rest Regularity Index), whereas others quantify multi-day stability by comparing each day to the individual’s average pattern (e.g., Interdaily Stability)^21^. Day-to-day metrics were designed to be more sensitive to insidious rhythm changes under the hypothesis that these insidious changes predispose individuals to circadian misalignment^22,23^. Evidence on day-to-day regularity and dementia risk is limited^17,18^, and its relation to cognitive decline has not been examined. These conceptual differences warrant systematic comparison to identify which aspects of regularity are most relevant for brain health.

Interpretation is complicated by the long prodromal phase of dementia, during which sleep-wake regularity itself may change as a consequence of neurodegenerative pathology and functional changes in mood, daily structure, and comorbidities^1,8,9^. These disease-related changes, such as destabilization of endogenous circadian rhythms or emergence of low-variability, uniform activity patterns that appear regular but are not health-promoting, may distort associations of sleep-wake regularity with dementia. These distortions may lead to life-course dependent associations: estimates in midlife may be less affected, whereas later-life associations may increasingly reflect emerging disease^24^. Beyond age, associations of sleep-wake disturbances with brain health may also vary by sex^25,26^ or APOE-ε4 status^10,27^, but this has not been systematically investigated for regularity.

To address these gaps, we conducted a two-step individual participant data (IPD) meta-analysis of 77,200 dementia-free middle-aged and older adults across six diverse prospective cohorts. We examined associations of three 24-hour sleep-wake regularity metrics with incident dementia and longitudinal trajectories of global cognition. Regularity was operationalized across three domains: day-to-day rest-wake regularity (Rest Regularity Index; RRI), day-to-day sleep-wake regularity (Sleep Regularity Index; SRI), and overall regularity of the activity rhythm (Interdaily Stability; IS), allowing direct comparison of three regularity constructs. Leveraging the large sample spanning midlife to late life, we further assessed whether associations differed by age, sex, and APOE-ε4 status.

## 2. Methods

### 2.1 Participants

We used data from six longitudinal cohorts across the United States and Europe: the Osteoporotic Fractures in Men Study (MrOS, sleep study between 2003-2005)^28,29^, the Rotterdam Study (RS, 2004-2007)^30^, The Multi-Ethnic Study of Atherosclerosis (MESA, 2010-2013)^31,32^, Rush Memory and Aging Project (MAP, 2005-2018)^33^, Whitehall II (WHII, 2012-2013)^34,35^, and the UK Biobank (UKB, 2013-2015)^36^. Detailed cohort descriptions are provided in Methods S1. Data were analyzed from June 2025 to March 2026. All participants provided written informed consent. Ethical approval was obtained from the institutional review boards of MrOS sites; Erasmus MC University Medical Center (RS); University of Washington (MESA); Rush University Medical Center (MAP); University College London (WHII); and National Information Governance Board for Health and Social Care and the NHS North West Centre for Research (UKB). We included all participants who completed actigraphy. Participants were excluded for lack of usable actigraphy data due to device malfunctioning or loss, insufficient valid recording days to calculate at least one regularity index (see below), prevalent dementia at actigraphy, or lack of follow-up for incident dementia (Figure S1). Shift workers were excluded if data were available (UKB, MESA), and UKB participants aged <55 years were excluded to minimize inclusion of early-onset dementia.

### 2.2 Actigraphy data collection

Participants wore wrist actigraphs (one-axial in MrOS, RS, MESA, MAP; tri-axial in WHII, UKB) to assess 24-hour sleep-wake patterns; median recording ranged from 4 (MrOS) to 10 days (MAP) (Methods S1). Raw signals were summarized at the epoch-level (activity counts or acceleration metrics), with non-wear periods excluded.

Three indices of 24-hour sleep-wake regularity were derived from actigraphy data: RRI, SRI, and IS (Figure 1). RRI and SRI measure day-to-day regularity, calculated as the probability of an individual being in the same state (rest/sleep vs. wake) at equivalent time-points 24-hours apart, averaged across the study length^23^. Theoretical background and computational details are provided in Methods S2. Cohort-specific algorithms classified each valid epoch as ‘sleep’ or ‘wake’. To calculate RRI, “rest” intervals were defined as sustained sequences of sleep-scored epochs lasting ≥ 5-minute. To calculate SRI, “sleep” intervals were defined as all sleep during the main nocturnal sleep episode and longer daytime sleep episodes lasting ≥ 30 minutes. RRI and SRI range from 0 to 100, with higher scores indicating greater regularity. RRI or SRI required at least 3 pairs of subsequent days with valid actigraphy; SRI was not calculated in MAP (main sleep window not defined) or WHII (for practical reasons). IS quantifies stability of the 24-hour activity rhythm across multiple days, comparing each day’s activity pattern with the individual’s mean pattern, based on activity counts aggregated into one-hour bins (Methods S3)^19,21,37^. IS ranges from 0 to 1, with higher values indicating greater stability, and was calculated for participants with ≥4 valid actigraphy days^38^.

**Figure 1:**
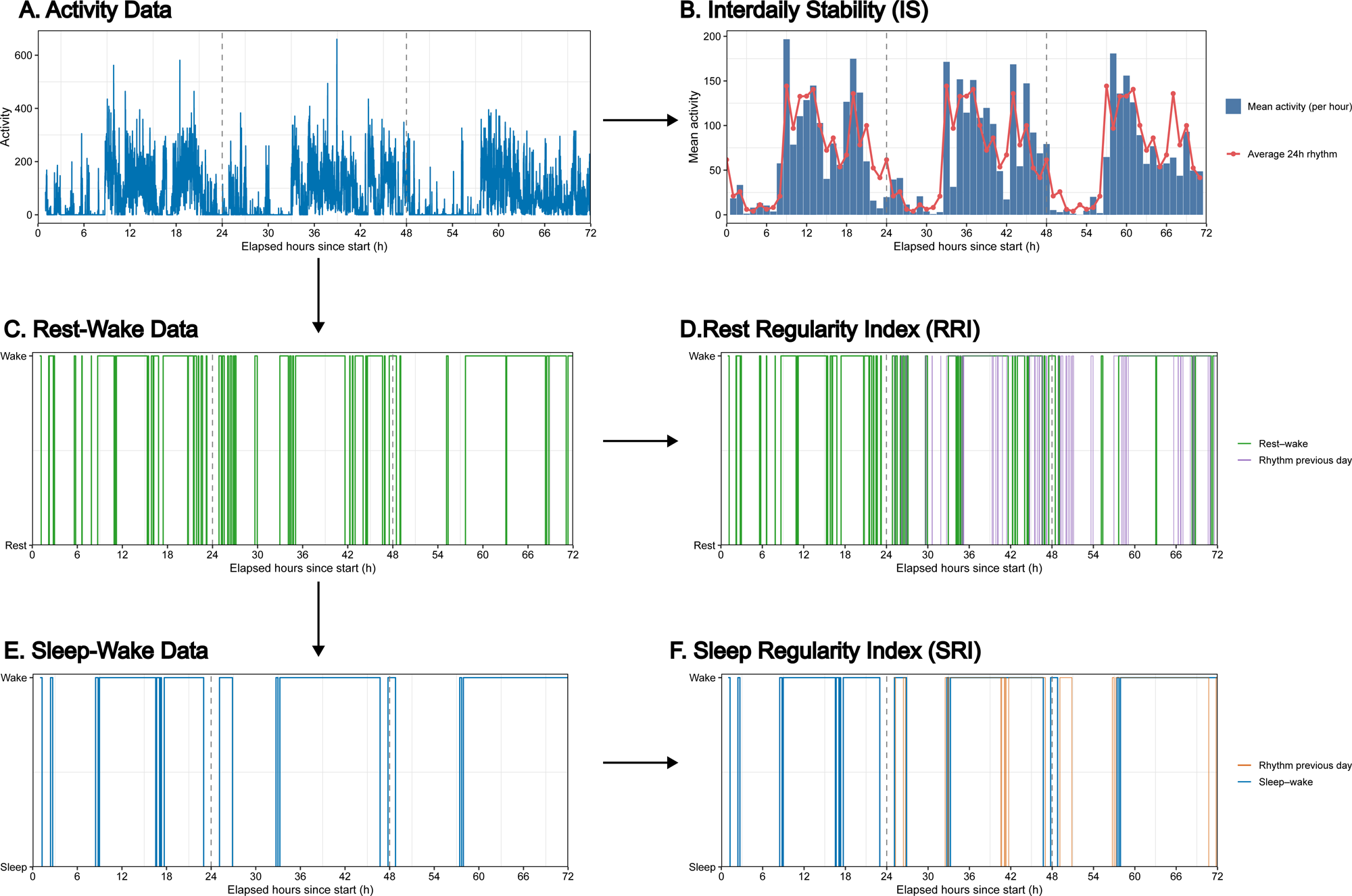
Schematic Visualization of different 24-hour sleep-wake regularity metrics. **Note:** The first 72 hours of recording are displayed for a randomly selected participant from MESA (Actiwatch Spectrum, Philips Respironics, Murrysville, PA). Activity values are summarized across 30 second epochs (A). Activity levels are summarized in hourly bins and Interdaily Stability (IS) is calculated by comparing hourly values with the average 24-hour activity pattern (B, red line). Cohort-specific algorithms classified each epoch as ‘sleep’ or ‘wake’; ‘rest’ intervals were defined as all ≥ 5-minute sustained sequences of sleep-scored epochs (C). Rest Regularity Index (RRI) is calculated by comparing each day’s rest-wake pattern to the previous day (D, purple line). ‘Sleep’ intervals are defined as all sleep during the nocturnal sleep episode and longer (≥ 30-minute) daytime sleep episodes (E). Sleep Regularity Index (SRI) is calculated by comparing each day’s sleep-wake pattern to the previous day (F, orange line).

### 2.3 Cognitive outcomes

Participants in all cohorts were followed for incident dementia using cohort-specific criteria (detailed in Methods S4). In MESA (mean follow-up: 9.0 years), WHII (6.9 years), and the UKB (7.9 years), dementia was identified through passive surveillance of hospitalization records, supplemented by death records in MESA and UKB^39–41^. In MESA, ascertainment was further strengthened by clinical adjudication at two visits. In RS (11.6 years), dementia was ascertained through active medical-record monitoring and routine cognitive screening at 4-yearly follow-up visits^42^. In MAP (7.2 years), diagnosis was based on annual cognitive testing with clinical adjudication^43^. In MrOS (7.1 years), probable dementia was assessed at follow-up visits based on self-reported physician diagnosis, dementia medication use, or significant decline in Modified Mini-Mental State (3MS) scores, defined as a decline ≥ 1.5 SD worse than the average change^44^. In four cohorts, global cognition was assessed repeatedly using cohort-specific instruments (3MS in MrOS^45^; a general cognitive factor in RS^46^; CASI in MESA^47^; and a composite of 19 tests in MAP^48^) and standardized as z-scores (Methods S5).

### 2.4 Covariates

Covariate data were collected at study entry or at the actigraphy visit; cohort-specific defnitions are in Methods S6. Covariates were primarily self-reported and included age, sex, race and ethnicity, study site, education, marital status, employment, body mass index, alcohol use, physical activity (actigraphy-based in UKB, self-report in other cohorts), smoking, cardiovascular comorbidities (stroke, heart attack, diabetes, and hypertension), depressive symptoms, and sleep medication use. In MrOS and MAP, missing covariate data were minimal and complete-case analysis was used; in MESA, RS, WHII, and UKB, missing data (<6% per covariate) were imputed using MissForest^49^.

### 2.5 Statistical analysis

We compared distributions, central tendencies, and correlations of regularity metrics, and quantified concordance with quartile transitions. Associations with age and sex were tested with Spearman correlations and Wilcoxon Rank-Sum tests. Cox-proportional hazards models assessed associations of sleep-wake regularity with incident dementia; proportional hazard assumptions were satisfied. Participants were censored at death, loss to follow-up, or study end. Non-linearity was evaluated using restricted cubic splines (10^th^, 50^th^, and 90^th^ percentiles); given minimal non-linearity, regularity was modeled linearly, with spline models retained as sensitivity analyses. Linear mixed effects models assessed associations with cognitive change over time. Models included random intercepts and interactions of follow-up time with regularity, age, sex, ethnicity and race, study site, and education, to account for potential steeper decline in cognitive performance across demographic groups. Random slopes were added if they significantly improved model fit (Likelihood Ratio Test).

Analyses were conducted separately by cohort and metric, with regularity indices standardized within cohorts, using minimally adjusted (Model 1; age, sex, ethnicity and race, study site, and education) and a fully adjusted (Model 2; all above-mentioned covariates) models. Cohort-specific estimates were pooled using random-effects meta-analysis. Heterogeneity was assessed using Cochran’s Q and I^2^. Stratified analyses were performed by age (<65, 65-<75, 75-<85 or ≥ 85 years), sex, and APOE-ε4 status; meta-regression assessed effect modification. Statistical significance was defined as 2-sided *p* < 0.05. Sensitivity analyses included Fine-Gray subdistribution hazard models to evaluate effects on cumulative dementia incidence rather than cause-specific hazards^50^; exclusion of cohorts in which dementia was identified primarily through hospitalization records (i.e., WHII and UKB); and additional adjustment for sleep duration (in RS, MrOS, MESA, and UKB).

## 3. Results

### 3.1 Baseline characteristics

A total of 74,733 adults were included across six cohorts. Mean age at sleep assessment ranged from 62.2 (standard deviation [SD]: 9.1) years in RS to 80.9 (7.3) in MAP, and the proportion of women ranged from 0% in MrOS to 77% in MAP (Table 1). Average scores ranged between 50.3-65.3 for RRI, 70.5-84.3 for SRI, and 0.53-0.80 for IS (Table 1, Figure S2). RRI declined with age across all cohorts (Figure 2; Spearman R= –0.02 to –0.20). Correlations of age with SRI were heterogeneous; negative in UKB and MESA, positive in RS, and absent in MrOS. For IS, correlation was positive in MAP, RS, and UKB, negative in MrOS and absent in MESA and WHII. Across cohorts, IS was higher in women than in men, but no consistent sex differences were observed for RRI and SRI (Figure S3, Table S1). Correlations between RRI and SRI were high in MrOS, MESA, and UKB (R=0.72-0.79) and moderate in RS (R=0.44) (Table S2). IS showed moderate correlations with both RRI and SRI in all cohorts (R*=*0.35-0.67). Quartile transition matrices showed that less than half of participants remained in the same quartile across indices, with transitions more frequent amongst those in Q2 and Q3 (Table S3).

**Figure 2:**
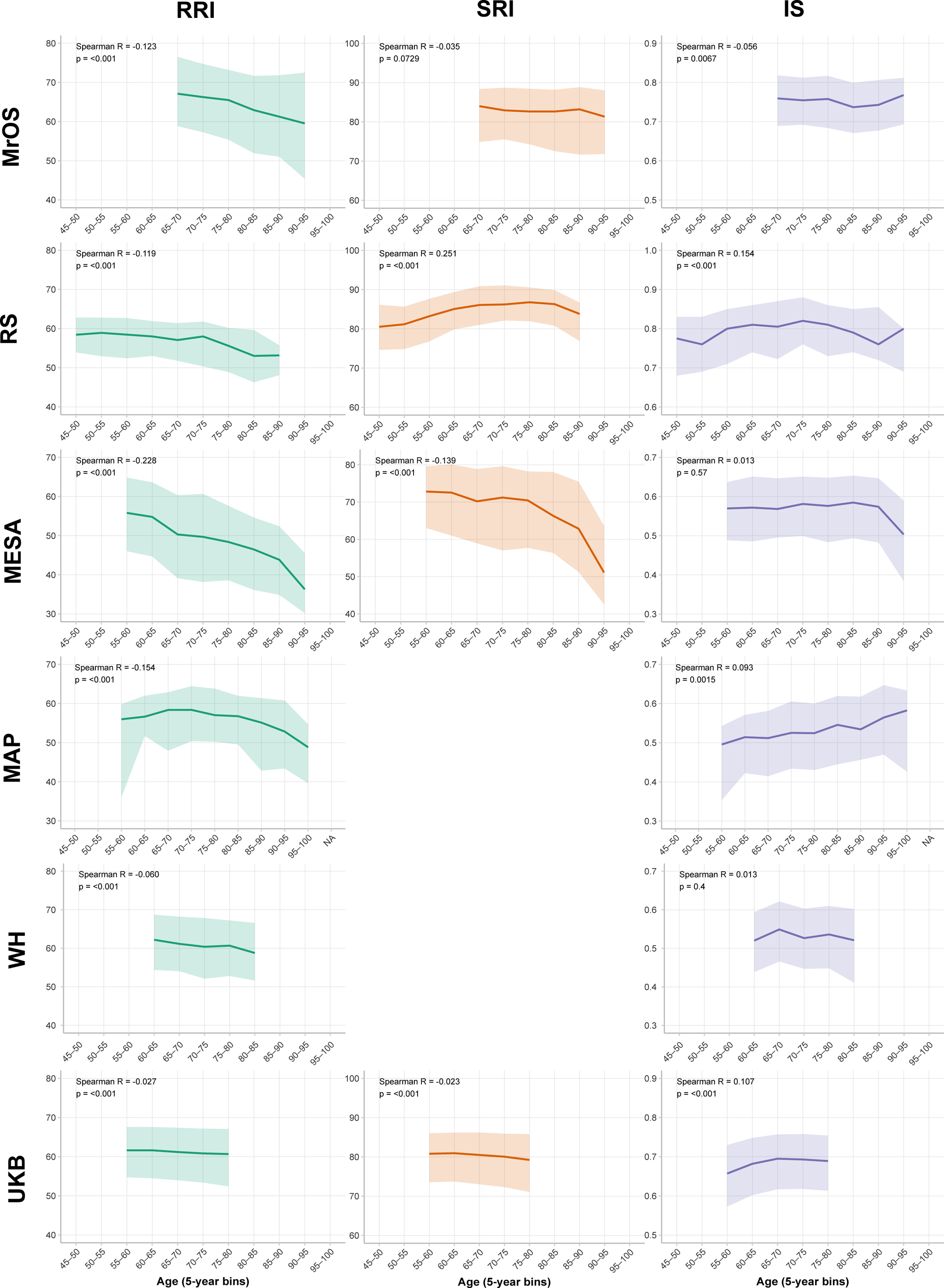
Correlations of 24-hour sleep-wake regularity with age across cohorts. **Note:** Age□related patterns in sleep□regularity metrics across all cohorts. Each panel displays median values of the Relative Regularity Index (RRI), Sleep Regularity Index (SRI), and Interdaily Stability (IS) within 5□year age bins, with shaded bands indicating the interquartile range. Values were summarized separately for each cohort to illustrate cross□sectional associations of age with each sleep□regularity metric. MrOS: Osteoporotic Fractures in Men Study; RS: Rotterdam Study; MESA: Multi-Ethnic Study of Atherosclerosis; MAP: Rush Memory and Aging Project; WHII: Whitehall II; UKB: UK Biobank.

**Table 1:**
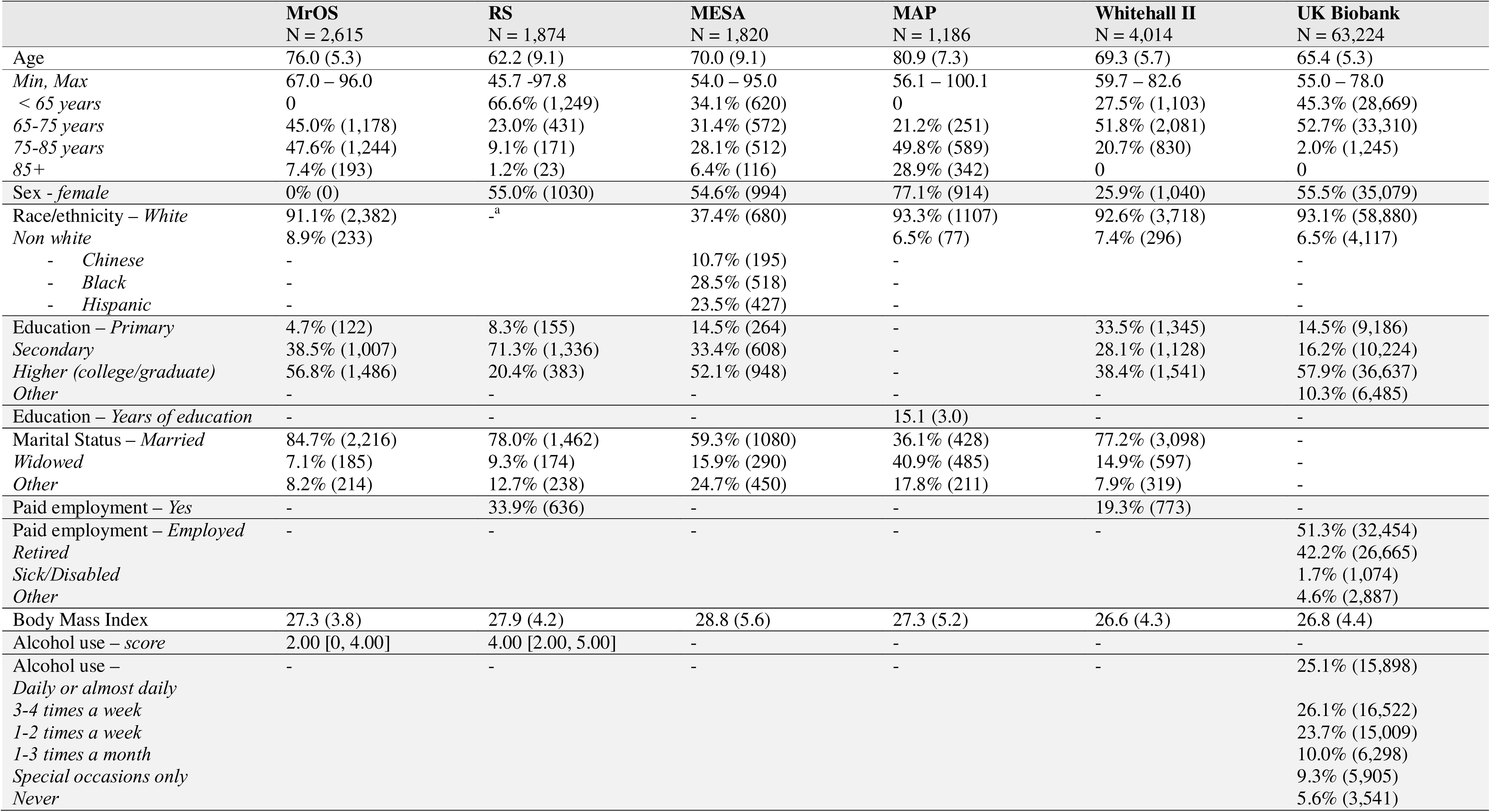
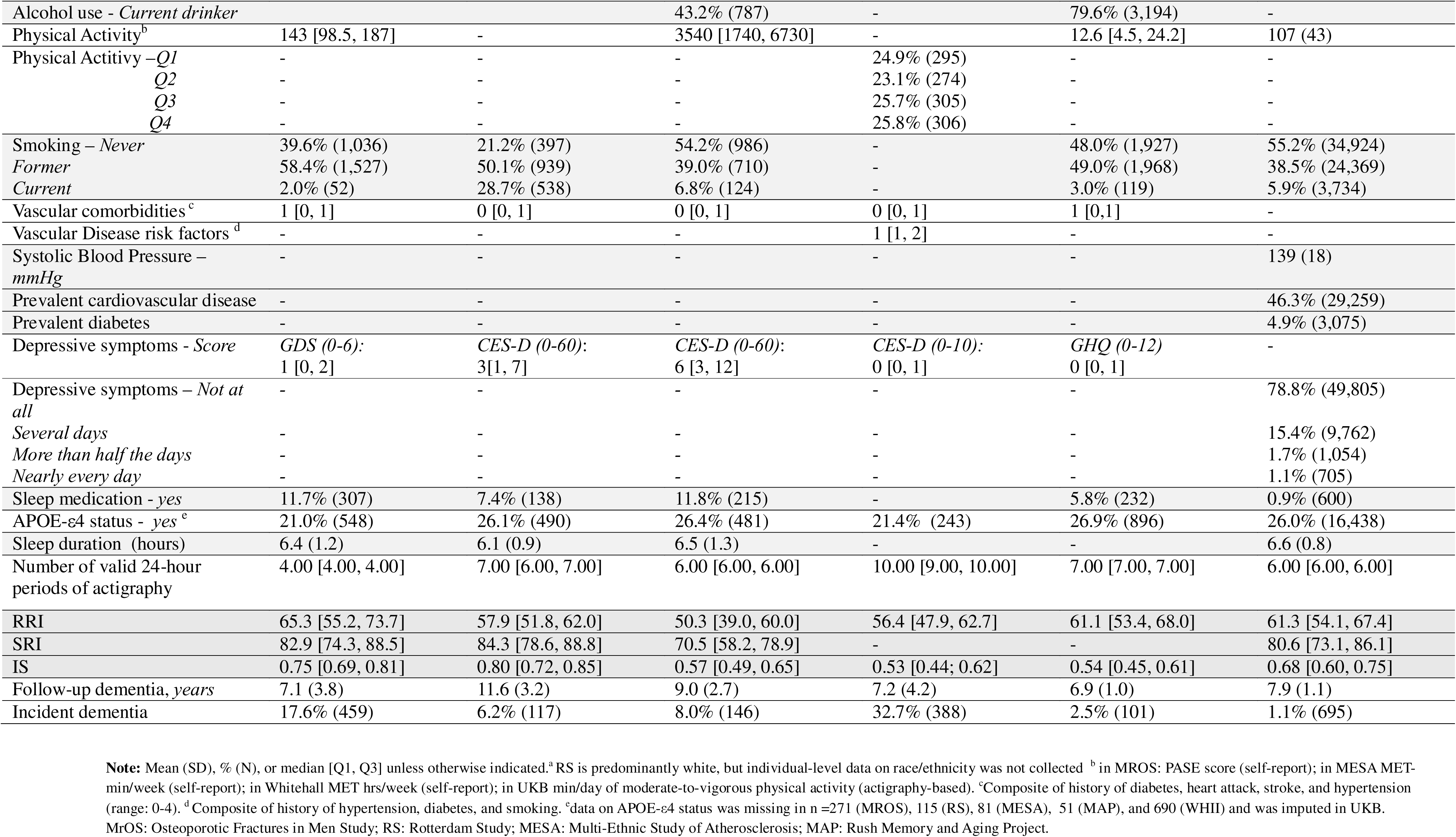
Sample Characteristics at baseline.

### 3.2 Dementia

Over a mean (SD) follow-up of 6.9 (1.0) to 11.6 (3.2) years, 1,888 incident dementia cases occurred. In minimally adjusted models (Model 1), greater regularity was associated with lower dementia risk across all three metrics (Table S4). After full covariate adjustment (Model 2), associations remained consistent: pooled hazard ratios (HRs) per 1-SD higher score were 0.86 (95%CI 0.79-0.95) for RRI, 0.87 (0.79-0.97) for SRI, and 0.91 (0.88-0.95) for IS (Figure 3, Table S5). For RRI, heterogeneity between cohorts was observed (*I*^2^:72.4%, *p*<0.01). Stratified analyses suggested stronger associations among middle-aged adults (e.g., pooled HR for RRI=0.69[0.49-0.96] for age<65 years) than older adults (e.g., pooled HR for RRI=0.85[0.71-1.02] for age>85 years), although meta-regression did not support a significant interaction with age (*p* > 0.55) (Figure S4). There was no evidence of effect modification by sex (*p >* 0.23, Figure S5) or APOE-ε4 status (*p* > 0.09, Figure S6). Spline models did not indicate substantial non-linearity (Figure S7). Effect sizes in Fine-Gray competing-risk models were smaller and not statistically significant for RRI and IS (Figure S8). Effect sizes were similar when cohorts relying on hospitalization-based dementia ascertainment (i.e. WHII and UKB) were excluded (Figure S9). Effect sizes were smaller and not statistically significant for RRI and IS when additionally adjusted for sleep duration (Figure S10).

**Figure 3:**
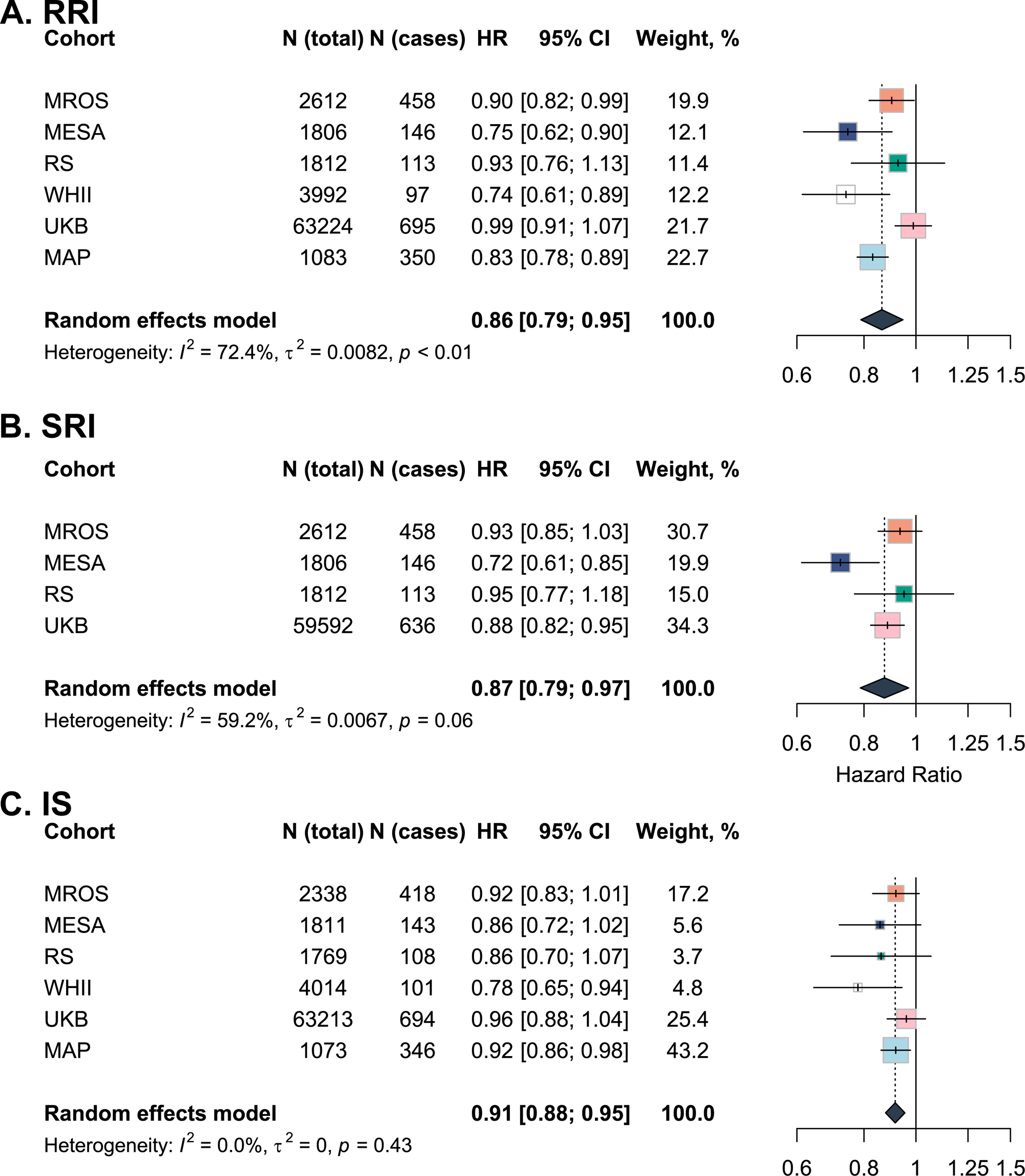
Association of 24-hour sleep-wake regularity with risk of all-cause dementia. **Note:** Effect sizes reflect hazard ratios with 95% confidence intervals per 1-SD higher regularity. Estimated with Cox Proportional Hazard models, adjusted for age, sex, race/ethnicity, study site, education, marital status, paid employment, body mass index, alcohol use, physical activity, smoking, vascular comorbidities, depressive symptoms, and use of sleep medication. Study-level estimates were pooled using random effects meta-analyses. RRI: Rest Regularity Index; SRI: Sleep Regularity Index; IS: Interdaily Stability; MrOS: Osteoporotic Fractures in Men Study; RS: Rotterdam Study; MESA: Multi-Ethnic Study of Atherosclerosis, WHII: Whitehall II; UKB: UK Biobank; MAP: Rush Memory and Ageing Project.

### 3.3 Cognitive Decline

Predicted trajectories of global cognition across quartiles of regularity are shown in Figure 4 for MESA as an example (other cohorts in Figures S11-13). Meta-analyzed results are presented in Figure 5 and Table S5. Higher SRI – but not RRI or IS – scores were associated with better global cognition at baseline (Table S5). Participants with higher RRI and IS had slower decline in global cognition over time, with pooled β*s* per 1-SD higher score per year of 0.003 (95%CI 0.001-0.006) for RRI and 0.003 (95%CI 0.000-0.006) for IS (Figure 5). Associations for SRI were similar in magnitude (β: 0.003; 95%CI, –0.002-0.008) but were not statistically significant and demonstrated substantial heterogeneity (*I*^2^:71.9%, *p*=0.03).

**Figure 4:**
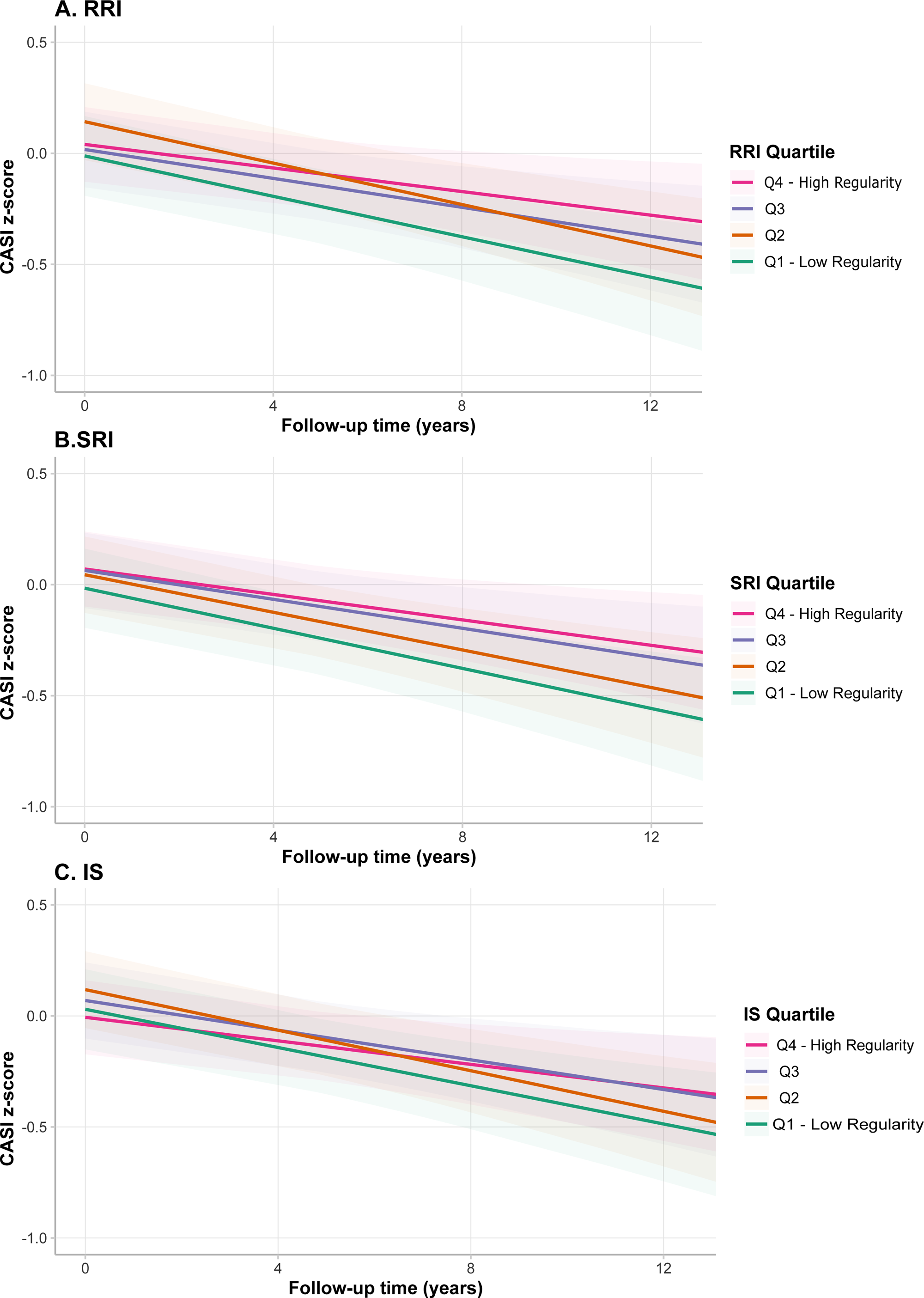
Predicted Cognitive Trajectories Across Quartiles of Regularity Metrics in MESA. **Note:** Visualizations for other cohorts are presented in Figures S11-S13. Panels A–C present model□based predicted trajectories of global cognitive performance (CASI z□scores) for the MESA cohort, estimated using linear mixed□effects models. Panel A shows trajectories across quartiles of the Relative Regularity Index (RRI), Panel B across quartiles of the Sleep Regularity Index (SRI), and Panel C across quartiles of the Interdaily Stability (IS) metric. Lines represent predicted values for each quartile (Q1–Q4), and shaded bands indicate 95% confidence intervals. All predictions are adjusted for age, sex, race/ethnicity, study site, education, marital status, paid employment, body mass index, alcohol use, physical activity, smoking, vascular comorbidities, depressive symptoms, and use of sleep medication.

**Figure 5:**
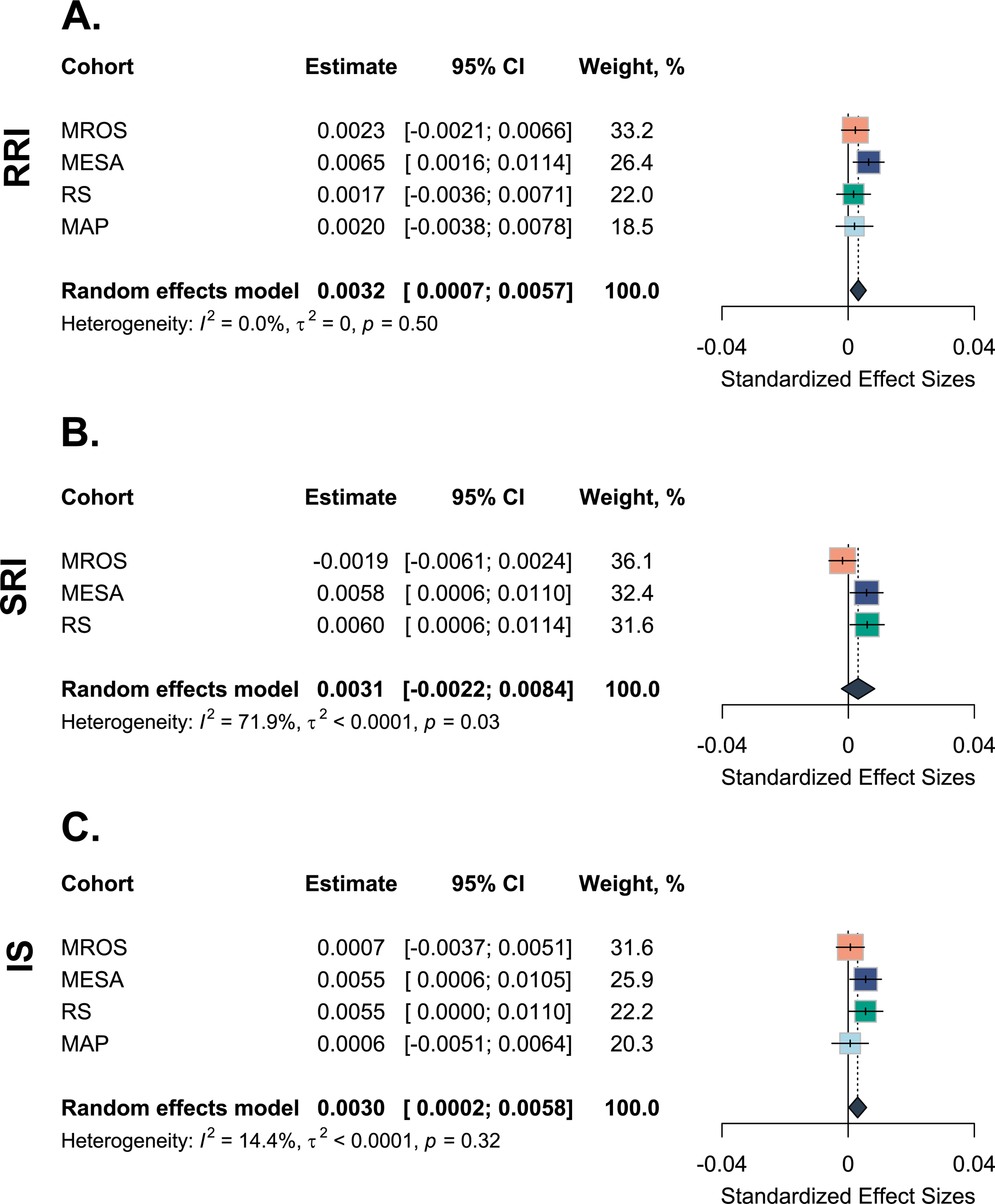
Association of sleep-wake regularity with global cognition over time. **Note:** All effect sizes are β[(95% CI] per 1-SD higher regularity per year. Models were adjusted for age, sex, race/ethnicity, study site, education, marital status, paid employment, body mass index, alcohol use, physical activity, smoking, vascular comorbidities, depressive symptoms, and use of sleep medication. Study-level estimates were pooled using random effects meta-analyses. Cognitive tests are standardized and if applicable transformed to improve normality. Higher scores indicate better cognitive performance. RRI: Rest Regularity Index; SRI: Sleep Regularity Index; IS: Interdaily Stability MrOS: Osteoporotic Fractures in Men Study; RS: Rotterdam Study; MESA: Multi-Ethnic Study of Atherosclerosis; MAP: Rush Memory and Aging Project. Follow-up time per cohort and main effects are noted in Table S5.

## 4. Discussion

In this two-step individual participant data meta-analysis of six longitudinal cohorts, greater regularity across rest-wake, sleep-wake, and 24-hour activity rhythms was associated with lower dementia risk and slower global cognitive decline. After accounting for demographics, health behaviors, depressive symptoms, and cardiovascular comorbidities, one-SD higher regularity was associated with an 9-14% lower risk of dementia. This indicates that maintaining regular 24-hour sleep-wake rhythms may be of interest for dementia prevention.

Most prior studies investigated regularity of 24-hour activity (IS) and reported mixed findings^10–12,15,16^. Lower day-to-day rest-wake (RRI) and sleep-wake (SRI) regularity were associated with higher dementia risk in two UKB studies^17,18^, but evidence outside this cohort was limited. By directly comparing three regularity constructs, we demonstrate that regularity across rest, sleep, and activity shows broadly similar associations with dementia risk and global cognition decline. Consistent with prior literature, cohort-specific effect sizes were heterogeneous and often modest. These nuanced effect sizes may have been difficult to detect in single-cohort studies, underscoring the value of a multi-cohort design. We further extend literature by demonstrating that irregular sleep-wake rhythms are associated with faster decline in global cognition, although effect sizes were small. Together, these findings suggest that subtle, everyday forms of irregularity are relevant to cognitive decline and dementia, extending previous evidence on shift work^5–7^. Randomized controlled trials are required determine whether these effects translate into clinically meaningful changes in cognitive health.

One important strength of our work is the direct comparison of three regularity constructs that differ in behavioral domain (sleep-wake, rest-wake, or activity patterns) and in whether they capture sequential day-to-day (RRI, SRI) regularity. Although correlations between RRI, SRI and IS were moderate to high, most participants were classified in different quartiles across metrics. This indicates only moderate concordance, in line with previous in-depth comparisons^23^. Across cohorts, RRI – but not SRI or IS – declined with age, whereas IS – but not RRI and SRI – was higher in women. The age-related decline of RRI may reflect its relative sensitivity to sleep fragmentation and brief daytime rest compared with other metrics. Overall, these patterns support that these metrics capture related but distinct dimensions of 24-hour sleep-wake regularity^23^. Despite conceptual differences, the similar associations with cognitive health suggests that metrics converge on a shared underlying regularity construct. Importantly, similar associations for SRI and IS underscore the relevance of 24-hour activity regularity beyond sleep.

Several complex and potentially bidirectional mechanisms may underlie these associations. Irregular 24-hour sleep-wake rhythms may promote circadian misalignment, reflecting misalignment between endogenous circadian rhythms and exogenous cues^51^, which may contribute to neurodegeneration through metabolic and endocrine dysregulation^4^, neuroinflammation, oxidative stress, and neuronal damage^52^. At the same time, irregularity may both contribute to and result from sleep disturbances, such as poor sleep quality and fragmentation, which may influence neurodegeneration^53,54^. Adjusiting for sleep duration attenuated effect sizes, suggesting that sleep-related mechanisms may partially underlie associations. Future mechanistic work incorporating multiple dimensions of sleep health is needed to disentangle these pathways. Neuroimaging studies further link irregular rhythms to reduced cortical thickness^55,56^, greater cerebrovascular burden^57,58^, and smaller temporal and hippocampal volumes^56^. By contrast, molecular biomarker evidence is less consistent, with mixed associations reported for imaging and plasma markers of AD-pathology and for plasma markers of neurodegeneration^27,52,59–64^. Similarly, effect sizes in Fine-Gray models were closer to null than primary analyses, which may reflect lower competing mortality among individuals with greater regularity^3^. Together, these findings support that irregular sleep-wake rhythms affect broader systemic mechanisms that influence both cognitive decline and mortality, rather than dementia-specific pathology alone.

The broad age range enabled examination of whether associations varied by age, an important consideration given potential reverse causation during the long prodromal phase of dementia. Across RRI, SRI, and IS, effect sizes were stronger among adults younger than 65 years and were smaller at older ages. This age-gradient may contribute to heterogeneity in prior studies, and suggests that reverse causation from prodromal dementia is unlikely to be the sole explanation, as such bias might be expected to strengthen associations at older age closer to dementia diagnosis. However, smaller effect sizes at older ages do not exclude reverse causation, because sleep-wake regularity in later life may also be affected by comorbidities, frailty, retirement, and functional limitations, potentially obscuring associations with dementia. Findings should be interpreted cautiously, as meta-regression did not support a statistically significant age interaction, and stratified analyses could not be adjusted for Model 2 covariates due to limited power and had less than 10 years of follow-up. Future studies beginning in midlife, with longer follow-up and repeated assessments of rhythm regularity and disease-progression markers, are needed to clarify temporal relationships.

### Strengths and Limitations

Major strengths include the harmonized calculation of three complementary 24-hour sleep-wake regularity indices across six prospective cohorts spanning midlife to late life. However, the two-step IPD meta-analytic approach has limited power to detect effect modification compared with fully pooled analyses^65^. Despite harmonized metric calculation, differences in actigraphy devices and processing likely contributed to between-cohort heterogeneity in distributions and correlations. Differences in dementia adjudication and cognitive testing may also contribute to heterogeneity, although results were similar when excluding cohorts relying on hospitalization-based dementia ascertainment. The observational study design limits causal inference, and unmeasured confounding and reverse causation may have biased effect estimates. Physical activity, which may both influence and be influenced by sleep-wake regularity, was not measured objectively in most cohorts, potentially resulting in residual confounding. Finally, regularity was measured once over a minimum of four days; although day-to-day metrics are relatively robust to shorter recordings^19^, longer or repeated measurements would better capture habitual patterns.

## 5. Conclusions

In this two-step IPD meta-analysis of six prospective cohorts, greater regularity in rest-wake, sleep-wake, and 24-hour activity rhythms were all associated with a lower risk of dementia and slower decline in global cognition. Further research is needed to evaluate whether highly irregular sleep-wake rhythms could serve as scalable markers for identifying individuals at risk for dementia, and randomized controlled trials are required to determine whether improving regularity may benefit long-term cognitive aging outcomes.

## Data Availability

Rotterdam Study: Data can be obtained upon request. Requests should be directed towards the management team of the Rotterdam Study, which has a protocol for approving data requests. MAP: Raw data are available by request through the Rush Alzheimer's Disease Center (RADC) Research Resource Sharing Hub https://www.radc.rush.edu/. Whitehall II data cannot be shared publicly because of constraints dictated by the study ethics approval and IRB restrictions. The Whitehall II data are available for sharing within the scientific community. Researchers can apply for data access at https://portal.dementiasplatform.uk/. UKB Data can be accessed following an approved application, which can be completed via the UKB website: [https://www.ukbiobank.ac.uk/].

## Acknowledgments

We thank all of the participating civil service departments and their welfare, personnel, and establishment officers; the British Occupational Health and Safety Agency; the British Council of Civil Service Unions; all participating civil servants in the Whitehall II study; and all members of the Whitehall II study team. The Whitehall II Study team comprises research scientists, statisticians, study coordinators, nurses, data managers, administrative assistants and data entry staff, who make the study possible.

We thank the MAP participants and their family members, and RADC faculty and staff.

This research has been conducted using the UK Biobank Resource under Application Number 70607. We thank the participants for volunteering their time and the UKB for making data and resources available.

## Data access, Responsibility, and Analysis

SJW Hoepel had full access to all the data in the study and takes responsibility for the integrity of the data and the accuracy of the data analysis. Authors SJW Hoepel, L Cribb and P Li conducted and are responsible for the data analysis.

## Data Sharing Statement

Rotterdam Study: Data can be obtained upon request. Requests should be directed towards the management team of the Rotterdam Study, which has a protocol for approving data requests. MAP: Raw data are available by request through the Rush Alzheimer’s Disease Center (RADC) Research Resource Sharing Hub https://www.radc.rush.edu/. Whitehall II data cannot be shared publicly because of constraints dictated by the study’s ethics approval and IRB restrictions. The Whitehall II data are available for sharing within the scientific community. Researchers can apply for data access at https://portal.dementiasplatform.uk/. UKB Data can be accessed following an approved application, which can be completed via the UKB website: [https://www.ukbiobank.ac.uk/].

## The Osteoporotic Fractures in Men (MrOS)

Study is supported by National Institutes of Health funding. The following institutes provide support: the National Institute on Aging (NIA), the National Institute of Arthritis and Musculoskeletal and Skin Diseases (NIAMS), the National Center for Advancing Translational Sciences (NCATS), and NIH Roadmap for Medical Research under the following grant numbers: U01 AG027810, U01 AG042124, U01 AG042139, U01 AG042140, U01 AG042143, U01 AG042145, U01 AG042168, U01 AR066160, R01 AG066671, and UL1 TR002369. The National Heart, Lung, and Blood Institute (NHLBI) provides funding for the MrOS Sleep ancillary study “Outcomes of Sleep Disorders in Older Men” under the following grant numbers: R01 HL071194, R01 HL070848, R01 HL070847, R01 HL070842, R01 HL070841, R01 HL070837, R01 HL070838, and R01 HL070839.The **Rotterdam Study (RS)** is funded by Erasmus Medical Center and Erasmus University, Rotterdam, Netherlands Organization for the Health Research and Development (ZonMw), the Research Institute for Diseases in the Elderly (RIDE), the Ministry of Education, Culture and Science, the Ministry for Health, Welfare and Sports, the European Commission (DG XII), and the Municipality of Rotterdam. **The Multi-Ethnic Study of Atherosclerosis (MESA)** Sleep Ancillary studies were funded by NIH-NHLBI R01HL098433 and NIH-NIA 5R01AG070867. The parent MESA study is supported by NHLBI funded contracts HHSN268201500003I, N01HC95159, N01HC95160, N01HC95161, N01HC95162, N01HC95163, N01HC95164, N01HC95165, N01HC95166, N01HC95167, N01HC95168 and N01HC95169, and by cooperative agreements UL1TR000040, UL1TR001079, and UL1TR001420 funded by NCATS. Cognitive data collection was supported by R01HL127659 from the National Heart, Lung, and Blood Institute, and grants R01AG054069 and R01AG058969 from the National Institute on Aging. A full list of participating MESA investigators and institutions can be found at http://www.mesanhlbi.org. **Rush Memory and Aging Project (MAP)** is funded under R01AG17917.The **Whitehall II (WHII)** study has been supported by grants from the National Institute on Aging, NIH (R01AG056477, R01AG062553); UK Medical Research Council (R024227, S011676), and the Wellcome Trust (221854/Z/20/Z).

