## Supplementary Material for "24-hour sleep-wake regularity and cognitive aging among 74,733 middle-aged and older adults from the US and Europe: The LifeSPAN Consortium"

### Supplementary methods 1: Cohort description & actigraphy data collection

A. **MrOS**

The MrOS Study was originally designed to assess risk factors for osteoporotic fractures in community-dwelling older men. MrOS is a multi-site cohort study including participants aged 65 and older from Birmingham Alabama, Minneapolis Minnesota, Palo Alto California, Pittsburgh PA, Portland Oregon, and San Diego California [1, 2] The baseline for the current study is the MrOS Sleep study, which is a sleep-focused substudy within the larger MrOS study, which took place between 2003 and 2005.

The SleepWatch-O (Ambulatory Monitoring, Inc, Ardsley, NY) was used to collect data on 24-hour activity. Participants were instructed to wear the actigraph continuously on their non-dominant wrist for a minimum of 5 consecutive 24-hour periods, except for during water-based activities. The SleepWatch-O measures movement with a piezoelectric linear accelerometer, which generated a voltage each time the actigraphy is moved. These voltages are summarized over 1-minute epochs in the proportional integration mode (PIM). To account for first-day effects, the data that was collected during daytime on the first day were removed. Thus, for each participant the data starts at the time participants got in-bed the first day they wore the watch. All 1-minute epochs in which the participant did not wear the actigraph were flagged as invalid.

B. **Rotterdam Study**

The Rotterdam Study (RS) is an ongoing prospective cohort of middle-aged and elderly persons in Rotterdam, the Netherlands and was designed to investigate determinants and consequences of aging and age-related disease [3]. All residents of Ommoord, a suburb of Rotterdam, aged 55 years and over were invited to participate in 1990 (cohort RS-I). In 2000 all residents aged 55 years and over, who had become 55 years of age or did not previously live in the study district, were recruited into the cohort (cohort RS-II). In 2006, all residents that were aged 45 years and older were additionally included into the cohort (RS-III).

The baseline of the current study is between 2004 and 2007, when participants were asked to wear an actigraph for 7 days and keep a sleep diary. Participants were instructed to wear the watch (Actiwatch model AW4, Cambridge Technology Ltd) continuously on their non-dominant wrist and remove it only when performing water-based activities. Additionally, participants pressed a marker button on the actigraph denoting bedtime and getting-up time. Non-wear time was identified as periods of at least 3 hours of consecutive zeroes.

C. **MESA**

The Multi-Ethnic Study of Atherosclerosis (MESA) is a prospective multi-site cohort study, designed to investigate the prevalence and progression of subclinical cardiovascular disease and to identify risk factors for cardiovascular disease in racially/ethnically diverse sample [4]. At study baseline (2000-2002) a total of 6,814 adults aged 45-84 who were free of clinically apparent cardiovascular disease at baseline were included. Participants were recruited from six US communities: Baltimore City and Baltimore County, MD; Chicago, IL; Forsyth County, NC; Los Angeles County, CA; Northern Manhattan and the Bronx, NY; and St. Paul, MN. The baseline of the current study is the MESA Sleep Ancillary Study, which took place around the time of MESA Exam 5 (2010-2013) [5]. Participants who reported regular use of oral devices, nocturnal oxygen, or nightly positive airway pressure devices were invited to participate.

Participants were instructed to wear the Actiwatch Spectrum (Philips Respironics, Murrysville, PA) on their non-dominant wrists for 7 consecutive days and completed a sleep diary during the same period. In addition, participants pressed an event marker at the time of sleep onset and offset. Actigraphy count data was summarized in 30 second epochs.

D. **MAP**

The Rush Memory and Aging Project (MAP) is a prospective observational study of adults over 65 recruited from the Chicago and broader northeastern Illinois Area in the United States, which has been ongoing since 1997. MAP is conducted at the Rush Alzheimer’s Disease Center and was designed to study cognitive decline and risk of Alzheimer’s disease and other health outcomes [6]. Participants enroll without known dementia and agree to annual evaluation and organ donation at death. It was approved by an Institutional Review Board of Rush University Medical Center and all participants signed informed and repository consents and an Anatomic Gift Act. Extensive clinical evaluations are repeated annually; actigraphy was introduced in 2005 to monitor motor actigraphy [7]. The baseline of the current study is for each participant the first annual visit in which they wore an actigraph.

Participants were instructed to wear the Actical device for 10 consecutive days on their non-dominant wrist at the time of the annual visit [7]. The Actical device measures accelerations primarily along one axis parallel to the face of the device with a continuous 32 Hz sampling frequency. Acceleration data is integrated into activity counts every 15 seconds. All periods of two hours or longer of consecutive epochs scored as sleep and with continuous low (<10) activity counts were regarded as non-wear time and excluded. During daytime, sequences of zeros with duration >60 minutes were regarded as non-wear time and excluded.

E. **Whitehall II**

The Whitehall II study is an ongoing prospective cohort study that was established in 1985-1988 among 10308 British civil servants [8]. Clinical examinations every four-five years since baseline. During the examination in 2012-2013 actigraphy was added to the data collection in all participants that were seen at the London clinic or living in the south-eastern regions of England and underwent clinical examination at home [9].

Participants were asked to wear a tri-axial accelerometer (GENEActiv Original; Activinsights Ltd, Kimbolton, UK) for nine consecutive days on their non-dominant wrist. Activity data was sampled at 85.7Hz and expressed relative to gravity. Non-wear time was detected using an algorithm that has been previously described [10].

F. **UK Biobank**

The UK Biobank is a large prospective cohort study of over 500,000 UK adults aged 40-69 years at baseline and is described in detail elsewhere [11]. Participants were recruited from UK National Health Service patient registers and enrolled between 2006–2010. The study contains extensive data from participants, including questionnaires, physical measures, accelerometer, and health outcomes. Between February 2013 and December 2015, a random selection of 236,519 UK biobank participants were invited to participate in the actigraphy substudy, of whom 106,053 agreed. Participants were instructed to wear a wrist-based triaxial actigraph (Axivity AX3; Axitivity, Newcastle upon Tyne, United Kingdom) for 7 days and nights.

### Supplementary methods 2: Regularity Indices (RRI and SRI)

#### **Theoretical background**

Two indices of day-to-day regularity – the Rest Regularity Index (RRI) and Sleep Regularity Index (SRI) – were calculated based on epoch-level binary data. Several methods have been put forward in the literature to calculate regularity indices – often all referred to as ‘Sleep Regularity Index’. They are all based on binary (sleep vs. wake) at the epoch level, but differ in what is considered ‘sleep’. As identified by Czeisler et al. [12], the two most commonly used metrics were implemented in the *GGIR* R package, reflecting regularity of any interval rest, regardless of length or whether it is a nap or nighttime sleep [13] and in the *SleepReg* package, identifying naps and WASO selectively by applying a threshold. [14, 15]. Differences between these two measures have been discussed by Czeisler et al. [12]. In this work we refer to the GGIR approach as ‘Rest Regularity Index’ and the Sleepreg approach as ‘Sleep Regularity Index’. Here, we attempt to derive similar metrics across all cohorts, balancing harmonization with feasibility. In each cohort, we followed previously established pre-processing pipelines to derive epoch-level sleep-wake estimates (See: Data Processing). We used abovementioned open source packages (i.e. *GGIR* and *SleepReg*) to derive RRI and SRI estimates if feasible. In MrOS, MESA, and MAP we used custom code based on these packages, which is made available online (<https://doi.org/10.5281/zenodo.19062652> ). Below, we note our definition of RRI, SRI, and the equation used. Next, we detail for each cohort how the metrics were calculated.

**Definitions**

**The Rest Regularity Index (RRI)** follows the definition and calculation as implemented in the GGIR package. The RRI is calculated as the regularity index of all intervals identified by the algorithm as ‘sleep’ (or ‘sustained inactivity bouts’), without applying any rules to specifically identify naps or wake after sleep onset (WASO). Importantly, the algorithm used to identify sustained inactivity bouts in GGIR applies a minimum duration of 5 minutes to any SIB interval, while the algorithms used in some other cohorts in this study (MrOS, MESA, RS, MAP) do not apply such a minimum and thus identifies short (i.e. 30 second or 1 minute) bouts of ‘sleep’ or wake. This is why we smoothed short (< 5 minute) intervals in MrOS, MESA, RS, and MAP to reduce high-frequency noise and harmonize the approach across cohorts, as detailed below.

**The Sleep Regularity Index (SRI)** was defined following the methods as specified in the *SleepReg* package [15] in which naps are defined as any interval of 30 minutes or longer with at least 95% sustained inactivity outside the main sleep episode and WASO as any epoch of 30 minutes or longer without any sustained sleep inactivity. Essentially, this means that data are ‘smoothed’ i.e. WASO or napping episodes shorter than 30 minutes are not considered, in order to separate out effects of sleep regularity from effects of rhythm fragmentation and sleep efficiency.

**Equation:** Rest Regularity Index (RRI) and Sleep Regularity Index (SRI) scores were calculated using this Regularity Index (RI) equation:

$$RI= - 100+\frac{200}{N_{v}}\left( \sum_{i=1}^{N} \delta\left( x_{i}, x_{i+1} \right) \right)$$

$$\text{where } \delta\left( x_{i}, x_{i+1} \right)=\left\{ \begin{aligned} 1, &x_{i}=x_{i+1}, \\ 0, x_{i}\neq x_{i+1}, \end{aligned} V= \left\{ i \right| x_{i}\neq NA, x_{i+1}\neq NA\}, N_{v}=\left| V \right| \right.$$

Sleep-wake state is represented by $x_{i}=1$ for sleep or rest, $x_{i}=0$ for wake, and $x_{i}=NA$ for excluded epochs.$N$ denotes the total number of recorded epochs, and $N_{v}= \left| V \right|$ is the number of valid epoch-by-epoch comparisons, considering only pairs where $x_{i}\neq NA$ and $x_{i+1}\neq NA$.RImetric theoretically ranges from -100 to 100, with 0 reflecting a completely random pattern and 100 a pattern characterized by rest/sleep and wake occurring at the exact same times each day. In practice, scores < 0 occur seldomly.

An RI score of 100 represents perfectly regular day-to-day sleep-wake patterns, and an RI score of 0 represents random patterns. RRI and SRI are both calculated for each day-pair and then averaged across all day-pairs for which enough valid data was available.

**Data preprocessing across cohorts**

**MrOS:** To determine sleep from wake time for each 1-minute epoch, the University of California San Diego (UCSD) scoring algorithm was used, as integrated in the Action W-2 software [16]. This algorithm calculates a moving average, considering the activity levels immediately prior to and after the current minute. Additionally, participants completed a sleep diary, which was used in the editing of the data to determine when the participant was in bed and when the actigraph was not worn. Scoring reliability and concordance with polysomnography were previously demonstrated [17, 18].

**Rotterdam Study:** Epoch-level count data was processed with GGIR version 3.2.11, using functionality for externally derived epoch-level data. For each 30 second epoch, sleep and wakefulness were distinguished using a validated algorithm with a threshold of 20 counts [19]. Time in bed was based on marker-based bedtime and getting-up time; missing marker times were imputed with sleep diary data.

**MESA:** Each 30 second epoch was scored as sleep or wake using the Actiware-Sleep v. 5.59 analysis software (Mini Mitter Co. Inc.), with a validated algorithm that analyzes the epoch in the context of the surrounding 2-min time period [20]. The sleep period was manually identified based on the event marker, sleep diary, light levels, and activity counts.

**MAP:** A previously validated algorithm based on activity counts [16, 21] was used to distinguish sleep from wake at the epoch-level. The recordings were resampled into an epoch length of 1 min (i.e., by summing up every four points). Next, each epoch was identified as sleep or wake based on a weighted sum of the current epoch, four epochs preceding the current epoch, and two epochs following the current one. Last, five rescoring rules were applied to initial scores to obtain final scores for sleep/wake at epoch-level.

**Whitehall II:** Sleep was estimated using a validated algorithm, implemented with GGIR (version 3.1-7). This algorithm has been previously described and evaluated against polysomnography data [22].

**UK Biobank:** Sleep was estimated using a validated algorithm, implemented with GGIR (2.7-1). This algorithm has been previously described and evaluated against polysomnography data [22].

#### **Calculation across cohorts**

**MrOS, MAP, and MESA:** We calculated the RRI and SRI from epoch-level data in MrOS, MAP, and MESA using custom code that is published online (<https://doi.org/10.5281/zenodo.19062652> ). To calculate the RRI, segments shorter than 5 minutes were evaluated within a surrounding neighborhood defined by half the minimum window size, rounded up (e.g., 3 minutes for a 5-minute window). If the neighborhood contained more sleep than wake, the segment was classified as sleep, and vice versa; ties were left unchanged. Thus, all segments of wake or rest were at least 5 minutes. To calculate the SRI, we used information on time in bed to separate all epochs into those that occur in bed and those that occur outside of bed. Only sleep intervals outside time in bed that contain 95% sleep for at least 30 minutes were retained. Similarly, only wake intervals that occur during time in bed that were at least 30 minutes were retained. A day-pair was considered valid if at least 16 hours were valid i.e. not missing in both days of the day-pair. For RRI and SRI, epoch-level data was inspected for individuals with values < 20 or > 97. Data for one individual in MrOS and one individual in MESA was excluded upon inspection, as an implausible amount of sleep was identified. As no information (i.e. diary/marker) was available to annotate “time in bed”, we could not calculate the SRI in MAP.

**RS:** The RRI was calculated within GGIR as output parameter SRI1, by setting a smoothing window (*SRI1_smoothing_wsize_hrs)* of 5 minutes and a smoothing fraction (*SRI1_smoothing_frac)* of 1 to yield results comparable to other cohorts. The SRI was calculated within GGIR as output parameter SRI2, by setting the following parameters: *possible_nap_duration =* (30, 240); *possible_nap_window* = (0, 24); *SRI2_WASOmin* = 30. A day-pair was considered valid if at least 16 hours were valid i.e. not missing in both days of the day-pair. RRI and SRI were only calculated in those with at least 3 valid day-pairs. For RRI and SRI, epoch-level data was inspected for individuals with values < 20 or > 97.

**Whitehall:** RRI was calculated as the SleepRegularityIndex, implemented in GGIR version 3.1-7. A day-pair was considered valid if at least 16 hours were valid i.e. not missing in both days of the day-pair. SRI was not calculated for practical reasons.

**UK Biobank:** Calculation of the RRI and SRI in the UK Biobank are described in more detail elsewhere [12]. In short, RRI was calculated with the SleepRegularityIndex function as implemented in GGIR (version 2.7-1) [13]. SRI was calculated by additionally implementing the *sleepreg* R package (version 1.3.5) [15]. Participants were required to have at least 7200 valid epoch pairs per recording to be included.

### Supplementary Methods 3: Interdaily Stability

**Definition**

Interdaily stability (IS) scores were calculated as the ratio of the variance of the average 24-hour profile and the overall variance in the data, using the following equation:

$$IS= \frac{N \sum_{h=1}^{p} {(X_{h}-X_{m})}^{2}}{p \sum_{i=1}^{N} {(X_{i}-X_{m})}^{2}}$$

N corresponds to the total number of data items, p is the number of data items per day (i.e. 24), X_m_ is the average of all data, X_h_ corresponds to each hour of the mean profile, while X_i_ represents each given hour of raw data. An IS score of 1 represents a perfectly regular activity rhythm, and an IS score of 0 represents random patterns.

**Data processing and calculation across cohorts**

**MrOS:** We summed the total number of invalid epochs for each 24-hour (midnight-to-midnight) day and calculated the number of days for each participant with a maximum of 4 hours of missing data. Participants were required to have at least 4 valid 24-hour periods of actigraphy to calculate IS.

**Rotterdam Study:** Non-wear time was identified as periods of at least 3 hours of consecutive zeroes and the 24-hour period following these periods were removed to prevent time-of-day effects. Participants were required to have at least 4 valid 24-hour periods of actigraphy to calculate IS. Calculation of IS was implemented with the nParAct R package, in which IS is calculated based on activity counts summarized over bins of 1 hour [23].

**MESA:** Days with more than 1 hour of non-wear time during sleep or less than 10 hours of wear time during wakefulness were excluded, participants were required to have at least 4 valid 24-hour periods of actigraphy to calculate IS. The algorithm started at 7AM of each day. IS was calculated based on hourly bins.

**MAP:** Calculation of IS in MAP has been described previously [24]. In short, data were resampled from 15-s intervals to hourly intervals. Participants were required to have at least 7 days of recording; if data length was 7 days or longer the first 7 days of recording were used.

**Whitehall II**: The calculation of IS in the Whitehall II cohort has been described previously [25]. In short, the Euclidean norm minus one (ENMO) of raw acceleration was calculated and corrected for calibration error and non-wear time. Acceleration values were averaged over 60-second epochs. Following previous studies using wrist-worn raw acceleration a 40 mg cut-point was used to differentiate between rest and active periods.

**UK Biobank:** The IS in the UKB was calculated an experimental implementation of the IS algorthims described in [26, 27] in GGIR (version 2.7-1).

### Supplementary methods 4: Dementia case ascertainment

A. **MrOS**

In MrOS presence of clinically significant cognitive performance was determined during four follow-up visits (2005-2006; 2007-2009; 2009-2012; 2014-2016). Following previous work in MrOS [28], clinically significant cognitive impairment was based on: 1) self-report of physician-diagnosed dementia; 2) use of dementia medication (based on examination of pill bottles); or 3) having a change in 3MS score ≥ 1.5 standard deviation worse than the average change from baseline to any follow-up visit. Once a participant met one of the criteria for clinically significant impairment at any visit, this was considered the endpoint for follow-up.

B. **Rotterdam Study**

In RS, participants were screened for dementia during each follow-up visit. Screening was based on the Mini-Mental State Examination and the Geriatric Mental Schedule organic level. Those with a Mini-Mental State Examination score less than 26 or Geriatric Mental Schedule score of more than 0 underwent further investigation and informant interview, including the Cambridge Examination for Mental Disorders of the Elderly. Additionally, participants were continuously monitored for dementia through linkage with medical records from general practitioners, the regional institute for outpatient mental health care, and nursing homes. Additional medical information (e.g. clinical notes and neuro-imaging reports) was obtained from hospital records, if available. Potential cases were reviewed by research physicians and discussed in consensus meeting led by a consultant neurologist. The diagnosis of all-cause dementia was based on DSM-III-R criteria [29]. Follow-up was complete until January 1^st^ 2020.

C. **MESA**

In MESA, incident dementia was ascertained through International Classification of Diseases (ICD) Ninth and Tenth revisions codes in medical records for hospitalizations reported during follow-up interviews and dementia death certificates. A previous validation study found that this method had a true positive rate of at least 73% in MESA [30]. Follow-up was completed until January 1^st^ 2020. Passive surveillance was supplemented with detailed cognitive testing and adjudication at MIND visits (2019-21, 2022-24, 2024-26). Cognitive function was assessed with a neuropsychological test battery consisting of the the Cognitive Abilities Screening Instrument (CASI) [31]; the Digit Symbol Coding (DSC) test [32]; the Digit Span (DS) test forwards and backwards[32]; the National Alzheimer’s Coordinating Center Uniform Data Set (UDS v3) neuropsychological battery[33]; the Wide Range Achievement Test (WRAT) [34]; the Auditory-Verbal Learning Test (AVLT)[35] and the Quick Dementia Rating System (QDRS) [36]. An informant appointed by the participant completed the QDRS, the Neuropsychiatric Inventory Questionnaire (NPI)[37] and the Functional Activities Questionnaire (FAQ/FAS)[38]. Cognitive testing was available in English, Spanish, Mandarin, and Cantonese with testing methods differing slightly by language and mode of administration. Normed cognitive testing results, QDRS, NPI-Q, Geriatric Depression Scale, physical examination, and clinical data were reviewd by an adjudication committee consisting of an expert panel of 8-10 investigators. The 2011 NIA-AA guidelines for MCI and dementia were used, independent of biomarker data, to adjudicate cognitive status as ‘cannot classify’, ‘no impairment’, ‘mild cognitive impairment’, and ‘probably dementia’ [39, 40]. Participants with probable dementia were included as cases in this analysis with diagnosis date set at the date of cognitive testing.

D. **MAP**

In MAP, clinical diagnosis of cognitive status is made at every annual home visit based on a three stage process including computer scoring of cognitive tests, clinical judgment of cognitive impairment by a neuropsychologist, and a diagnostic classification of dementia by a clinician [41-43]. Clinical diagnosis of dementia and clinical Alzheimer's dementia are based on NINCDS/ADRDA criteria.

E. **Whitehall II**

In Whitehall, dementia cases were identified with ICD-10 codes F00, F03, F05.1, G30, and G31 through linkage to the national Hospital Episode Statistics (HES) database with unique National Health Service (NHS) identification numbers. Follow-up was performed up to March 31^st^ 2019. The NHS performs most of the out- and in-patient healthcare and identification of dementia cases in this dataset has previously been shown to be adequate (sensitivity: 78.0%; specificity: 92.0%) [44].

F. **UK Biobank**

In UKB, all-cause dementia cases were identified through linkage with primary care data, hospital admission data, and death registries. The accuracy of dementia case identification is high, with a positive predictive value >80% for each data source [45]. Follow-up was complete until January 1^st^ 2023.

### Supplementary methods 5: Repeatedly measured cognitive performance

A. **MrOS**

In MrOS, cognitive tests were administrated by trained staff to assess cognitive function at the time of actigraphy and at four follow-up visits (2005-2006; 2007-2009; 2009-2012; 2014-2016). The 3MS is a global measurement of cognition, with components for orientation, concentration, language, praxis, and immediate and delayed memory. Scores range from 0 to 100, with higher scores representing better cognitive function [46]. To account for ceiling effect, all 3MS scores were adjusted with the following equation: log3MS = -log10(101 – 3MS). Global cognition was measured by standardized transformed 3MS scores, using the mean and standard deviation from Sleep Visit 1.

B. **Rotterdam Study**

In the Rotterdam Study, cognition was assessed with a cognitive test battery at four follow-up (2009-2014; 2014-2016; 2018-2019; 2021-2024) visits at the research center. The test battery consisted of the Letter Digit Substitution Test, Stroop Interference Test, Word Fluency Test, 15-Word Learning test, and Purdue Pegboard Test and has been previously described in more detail [47]. In line with previous work in the Rotterdam Study, global cognition was operationalized with a principle component analysis to compute a standardized general cognitive factor (*g*-factor) in each round, using results from the five above-mentioned tests [48]. The first unrotated component comprised the g-factor and was converted into a standardized Z-score.

C. **MESA**

In MESA, cognition was measured in-person during Exam 5 (years 2010-2012), Exam 6 (years 2016-2018), and Exam 7 (years 2022-2023). The baseline for cognitive function was Exam 5; models were adjusted for the potential time-gap between the Sleep Visit and Exam 5. All findings were similar when 712 participants with a time-gap of > 1 years were excluded (results not shown). The Cognitive Abilities Screening Instrument (CASI) contains 25 items that reflect 9 cognitive domains. Individual items were summed to provide an overall cognitive function score (range 0-100) [31]. Cognitive tests from participants who had > 3 missing CASI components, a CASI score < 20 and/or were deemed as probable invalid for other reasons were not used. To account for ceiling effect, all CASI scores were adjusted with the following equation: logCASI = -log10(101 – CASI). Global cognition was measured with standardized logCASI scores using the mean and standard deviation from Exam 5.

D. **MAP**

In MAP, a comprehensive battery of 21 cognitive tests is performed during annual home visits [49]. Of these, 19 are used to generate a global cognition score calculated as the average of all standardized tests scores, if at least half of the raw scores for that visit were present.

### Supplementary methods 6: Measurement of covariates across cohorts

| Covariates | **MrOS** | **RS** | **MESA** | **MAP** | **Whitehall II** | **UK Biobank** |
| --- | --- | --- | --- | --- | --- | --- |
| **Age** | Age at sleep visit (years) | Age at sleep visit (years) | Age at sleep visit (years) | Age at sleep visit (years) | Age at sleep visit (years) | Age at sleep visit (years) |
| **Sex** | - Male | - Male - Female | - Male - Female | - Male - Female | - Male - Female | - Male - Female |
| **Race/ ethnicity** | - White - Non-White: African American/ Black, Asian, Pacific Islander, American Indian, Alaskan Native, Multiracial, Hispanic, Unknown | Not available | - White - Chinese - Black - Hispanic | - White - Non-White:  African American/ Black, Asian, Pacific Islander, American Indian, Alaskan Native, Other, Unknown | - White - Non-White | - White - Non-White |
| **Site** | Study site | Study subcohort:  RS-I, RS-II or RS-III | Study site | - | - | - |
| **Education** | - Primary: Elementary school or some high school (yrs 1-12) - Secondary: High school or some college (yrs 12-16) - College/ university/ graduate school: College or graduate school (yrs 16 or more) | - Primary*:* Primary Education - Secondary: general education or lower/intermediate vocational education - College/ university/ graduate school: Higher vocational or university | - Primary: No schooling or grades 1-11 - Secondary: High School, GED, or some college - College/ university/ graduate school: Technical School, Associate Degree, Bachelor’s Degree, Graduate or Professional School | Years of education | - Primary: Elementary or lower secondary - Secondary: Higher secondary - College/ university/ graduate school: Degree or higher | - Primary*:* GSE-GCSE or equivalent - Secondary: A-levels & NVQ or equivalent - College/ University/ Graduate school*:* College or university degree - Other |
| **Marital Status** | At time of sleep visit:   - Married Widowed - Other: Divorced, separated, never married, other | At time of sleep visit:   - Married Widowed - Other: Divorced, separated, single, never married, other | At time of sleep visit:   - Married: Married or living as married - Widowed - Other: Divorced, separated, never married, other | At study entry:   - Married - Widowed - Other: Never married, divorced, separated | At study entry:   - Married: Married or cohabitating - Single: Single - Other: Divorced, widowed | Not available |
| **Paid employment** | Not available | - Paid employment - No paid employment: Registered unemployed, housewife, not able to work, rentier, early retirement, retirement | Not available | Not available | - Paid employment: Civil service or non-civil service employment - No paid employment: not working, retired, out of work, long-time sickness | - Paid employment*:* in paid employment or self-employed - Retired - Sick/Disabled: Unable to work because of sickness or disability - Other: unemployed; looking after home/family; unpaid or voluntary work; full or part-time student; other |
| **Body mass index** | Body mass index (kg/m^2) | Body mass index (kg/m^2) | Body mass index (kg/m^2) | Body mass index (kg/m^2) | Body mass index (kg/m^2) | Body mass index (kg/m^2) |
| **Alcohol use** | Score [0-5]:  0: No drinks in the past 12 months  1: Less than one drink per week  2: 1-2 drinks per week  3: 3-5 drinks per week  4: 6-13 drinks per week  5: 14 or more drinks per week | Score [0-5]:  0: No drinks in the past 12 months  1: Less than one drink per week  2: 1-2 drinks per week  3: 3-5 drinks per week  4: 6-13 drinks per week  5: 14 or more drinks per week | Currently drinking alcohol:   - Yes - No | Not available | Alcohol use in the past week:   - Yes - No | Alcohol frequency:   - 1: Daily or almost daily - 2: 3-4 times a week - 3: 1-2 times a week - 4: 1-3 times a month - 5: Special occasions only - 6: Never |
| **Physical Activity** | PASE Score | not available | Moderate and vigorous physical activity:  *MET-*min/week | Self-reported hours per week in physical activity summed over 5 categories;  analyzed with quartiles following Rush conventions | MET hrs/week for moderate and vigorous activity | Moderate-to-vigorous physical activity (MVPA)  min/day – Actigraphy based |
| **Smoking** | - Never - Former - Current | - Never - Former - Current | - Never - Former - Current | Not available – included in vascular disease risk factors | - Never - Former - Current | - Never - Former - Current |
| **Cardio-vascular comorbidities** | Composite score (0-4) of stroke, heart attack, diabetes, and hypertension | Composite score (0-4) of stroke, heart attack, diabetes, and hypertension | Composite score (0-4) of stroke, heart attack, diabetes, and hypertension | Vascular disease burden:  Composite score (0-4) of claudication, stroke, heart conditions, and congestive heart failure  Vascular disease risk factors: Composite score (0-3) of hypertension, diabetes, and smoking history | Composite score (0-4) of stroke, heart attack, diabetes, and hypertension | Prevalent CVD:  Self-reported CVD at UKB baseline or a CVD diagnosis (ICD-10 codes I00-I99 and C00-C97) at the time of actigraphy.  Prevalent diabetes  Systolic BP (mmHG) |
| **Depressive symptoms** | Geriatric Depression Score (GDS)  Ranges 0-6 | Center for Epidemiologic Studies Depression scale (CES-D)  Ranges 0-30 | Center for Epidemiologic Studies Depression Scale (CES-D)  Ranges 0-30 | Center for Epidemiologic Studies Depression scale (CES-D10)  Ranges 0-10 | GHQ: Depression subscale  Ranges 0-12 | Over the last 2 weeks, how often have you been bothered by depressive symptoms?  Not at all  Several days  More than half the days  Nearly every day |
| **Use of sleep medication** | Self-report use of sleep medication in the past month   - Yes: At least once a week - No: Less than once a week | Self-report use of sleep medication in the past month   - Yes: At least once a week - No: Less than once a week | Use of sleeping pills in the past 4 weeks:   - Yes: At least once a week - No: Less than once a week | Not available | Use of hypnotics, anxiolytics, antipsychotics, or antidepressants:   - Yes - No | Use of insomnia medication at UKB baseline   - Yes - No |

### Literature Supplementary methods

20.	Oakley, N.R., *Validation with Polysomnography of the Sleepwatch Sleep/Wake Scoring Algorithm used by the Actiwatch Activity Monitoring System*. 1997, Cambridge Neurotechnology Ltd.

32.	Wechsler, D., *Wechsler adult intelligence scale.* Archives of Clinical Neuropsychology, 1955.

33.	Weintraub, S., et al., *Version 3 of the Alzheimer Disease Centers' Neuropsychological Test Battery in the Uniform Data Set (UDS).* Alzheimer disease and associated disorders, 2018. **32**(1): p. 10-17.

34.	Wilkinson, G. and G.J.R.C.B. Robertson, *Wide range achievement test 4 professional manual: psychological assessment resources.* 2006. **52**: p. 57-60.

35.	Schmidt, M., *Rey Auditory Verbal Learning Test : RAVLT : a handbook*. RAVLT : a handbook. 1996, Los Angeles, CA: Western Psychological Services.

### Supplementary Figure 1: Flowchart of the study sample

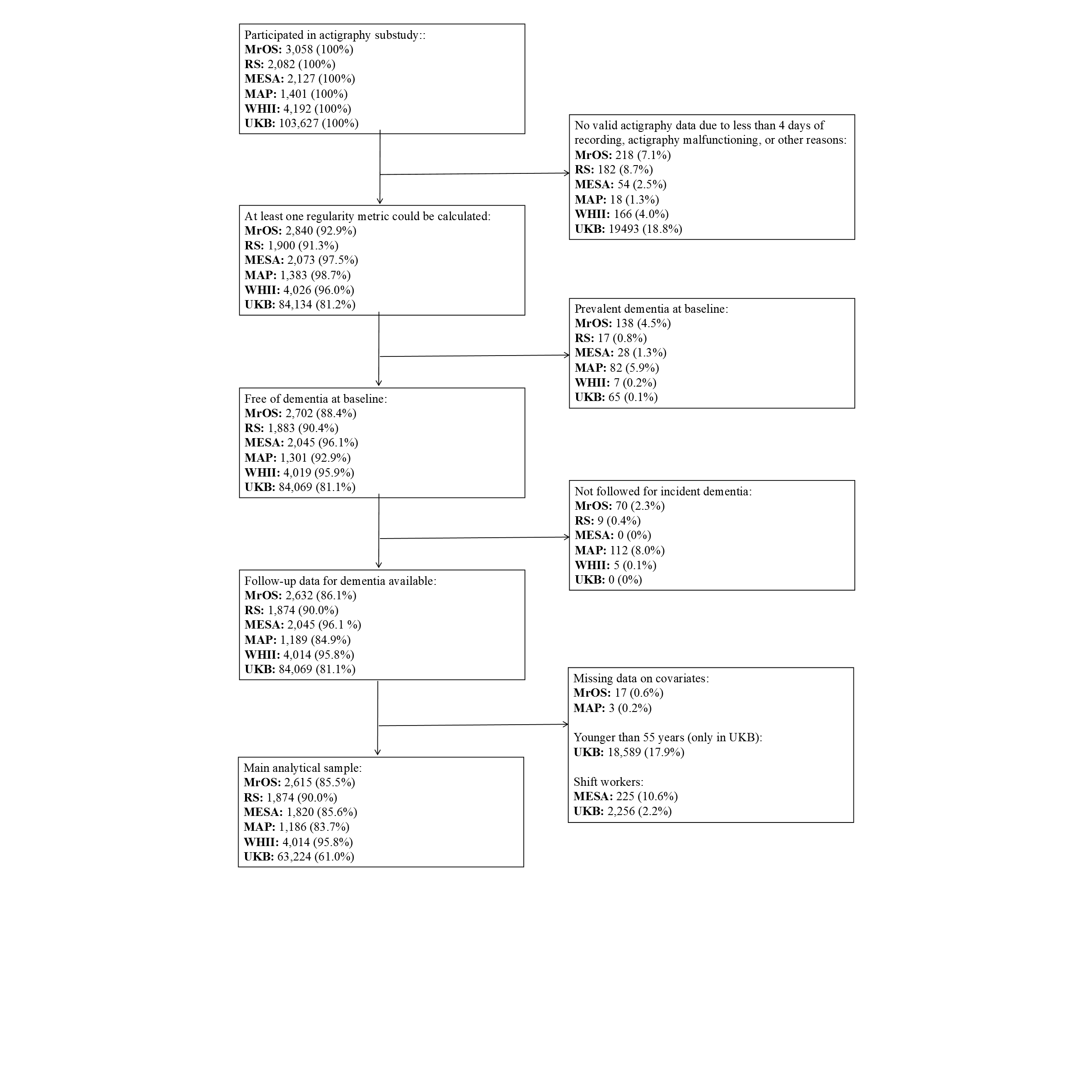

**Note:** MrOS: Osteoporotic Fractures in Men Study; RS: Rotterdam Study; MESA: Multi-Ethnic Study of Atherosclerosis; MAP: Rush Memory and Aging Project; WH: Whitehall II; UKB: UK Biobank.

Supplementary Figure 2: Distribution of Regularity Metrics across cohorts

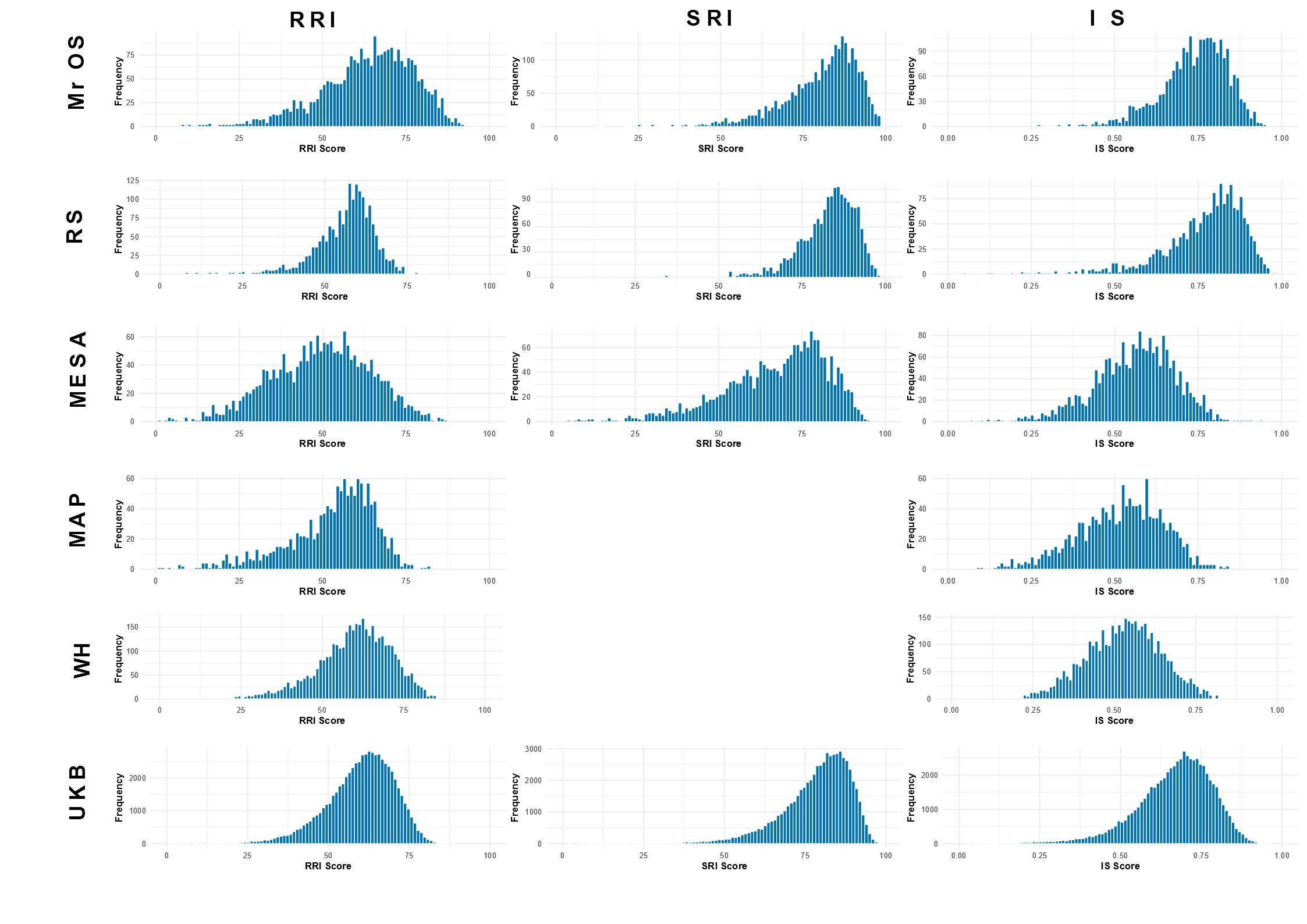

**Notes:** Histograms visualize distribution of Rest Regularity Index (RRI, practical range 0-100); Sleep Regularity Index (SRI, practical range 0-100); Interdaily Stability (IS, range 0-1) across cohorts. MrOS: Osteoporotic Fractures in Men Study; RS: Rotterdam Study; MESA: Multi-Ethnic Study of Atherosclerosis; MAP: Rush Memory and Aging Project. Bins reflecting 5 individuals or less are not displayed for WH to comply with cohort-specific privacy regulations.

### Supplementary Figure 3: Sex-differences in regularity

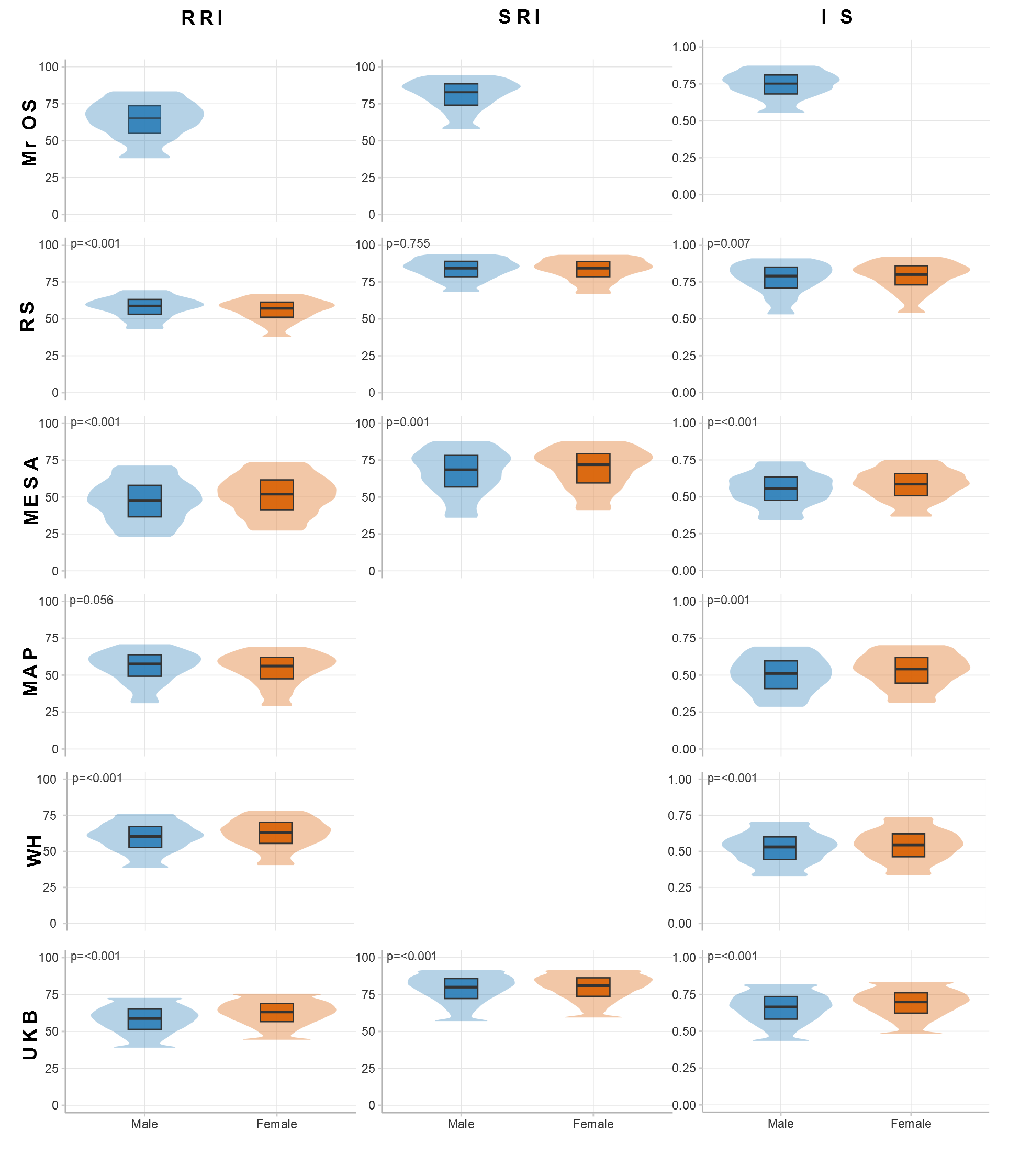

**Note:** Box-plots and violin plots visualize sex-differences in distribution of Rest Regularity Index (RRI), Sleep Regularity Index (SRI) and Interdaily Stability (IS) across cohorts. Violin plots show the 5^th^ – 95^th^ percentile of distribution to comply with privacy regulations. P-values for Wilcoxon Rank-Sum tests for differences between sexes are provided. MrOS: Osteoporotic Fractures in Men Study; RS: Rotterdam Study; MESA: Multi-Ethnic Study of Atherosclerosis; MAP: Rush Memory and Aging Project; WH: Whitehall II; UKB: UK Biobank.

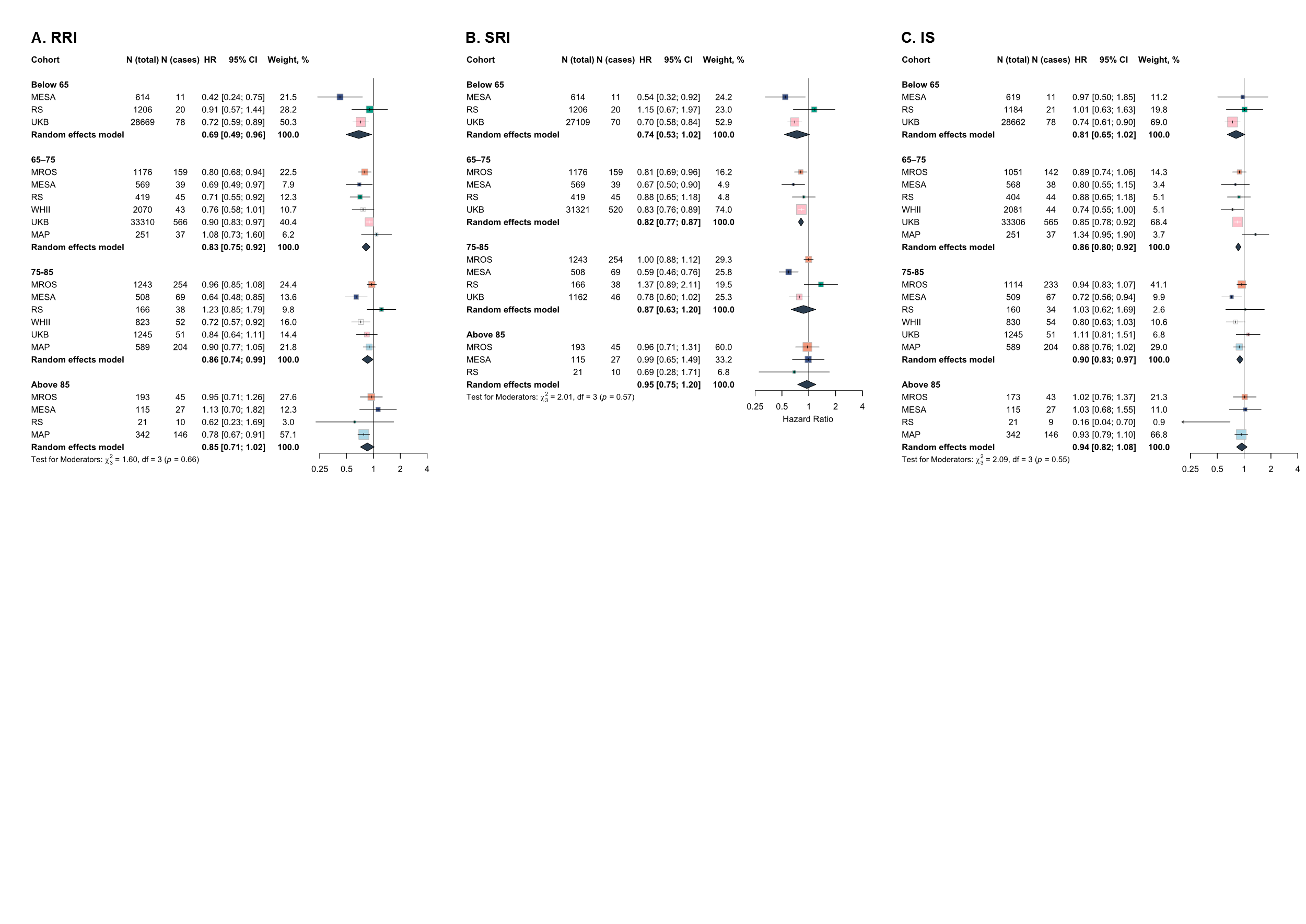
Supplementary Figure 4: Age-stratified association of 24-hour sleep-wake regularity with risk of all-cause dementia

**Note:** Effect sizes reflect hazard ratios with 95% confidence intervals per one-SD higher regularity score. Estimated with Cox Proportional Hazard models, adjusted for age, sex, race andethnicity, study site, and education. Study-level estimates were pooled using random effects meta-analyses and meta-regression was conducted to explore the moderating effects of age. RRI: Rest Regularity Index; SRI: Sleep Regularity Index; IS: Interdaily Stability; MrOS: Osteoporotic Fractures in Men Study; RS: Rotterdam Study; MESA: Multi-Ethnic Study of Atherosclerosis; WH: Whitehall II; UKB: UK Biobank; MAP: Rush Memory and Ageing Project. Follow-up length for these analyses is presented in Supplementary Table S4.

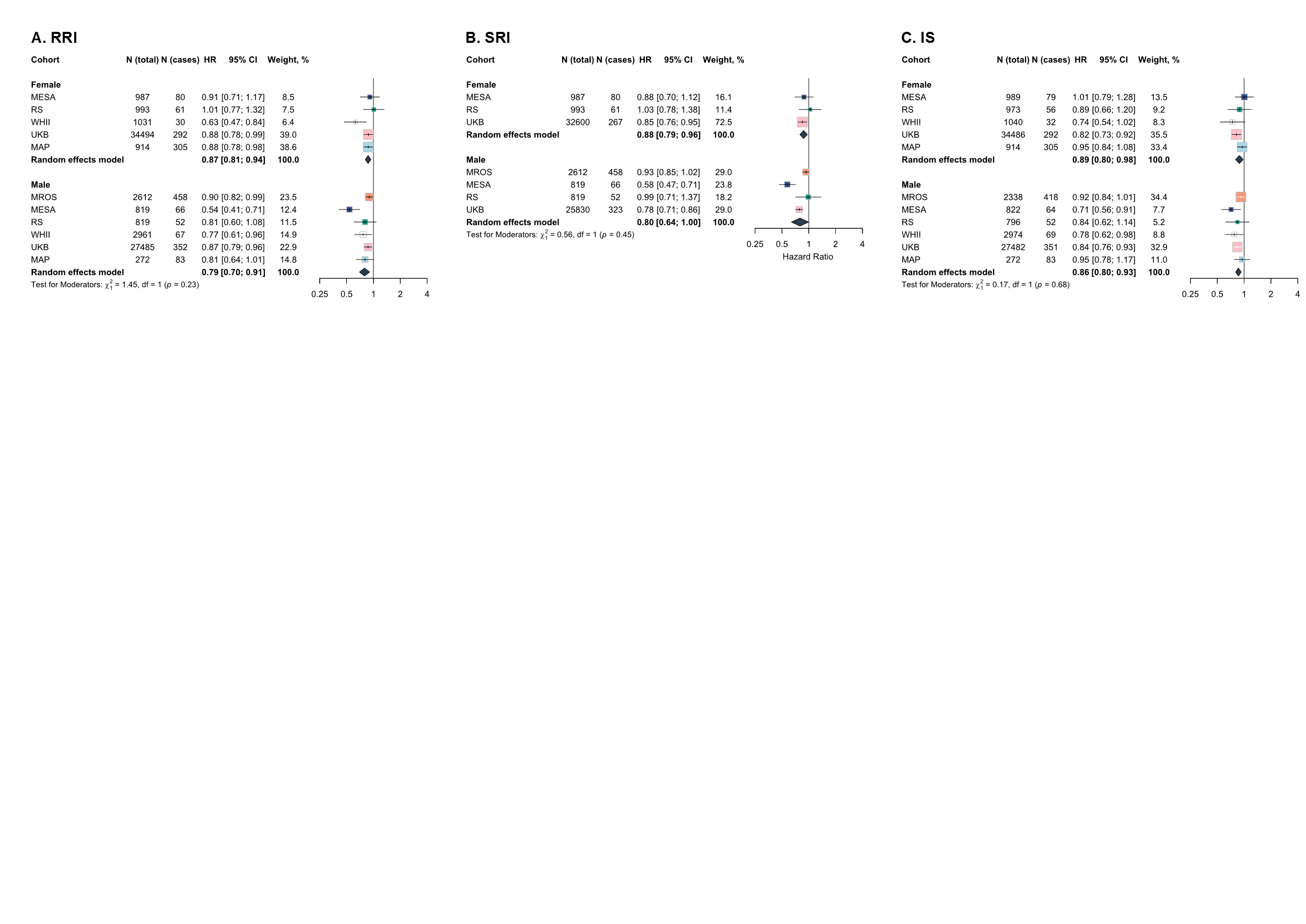
Supplementary Figure 5: Sex-stratified association of 24-hour sleep-wake regularity with risk of all-cause dementia

**Note:** Effect sizes reflect hazard ratios with 95% confidence intervals per on SD higher regularity score. Estimated with Cox Proportional Hazard models, adjusted for age, sex, race/ethnicity, study site, and education. Study-level estimates were pooled using random effects meta-analyses and meta-regression was conducted to explore the moderating effects of sex. RRI: Rest Regularity Index; SRI: Sleep Regularity Index; IS: Interdaily Stability; MrOS: Osteoporotic Fractures in Men Study; RS: Rotterdam Study; MESA: Multi-Ethnic Study of Atherosclerosis; WH: Whitehall II; UKB: UK Biobank; MAP: Rush Memory and Ageing Project.

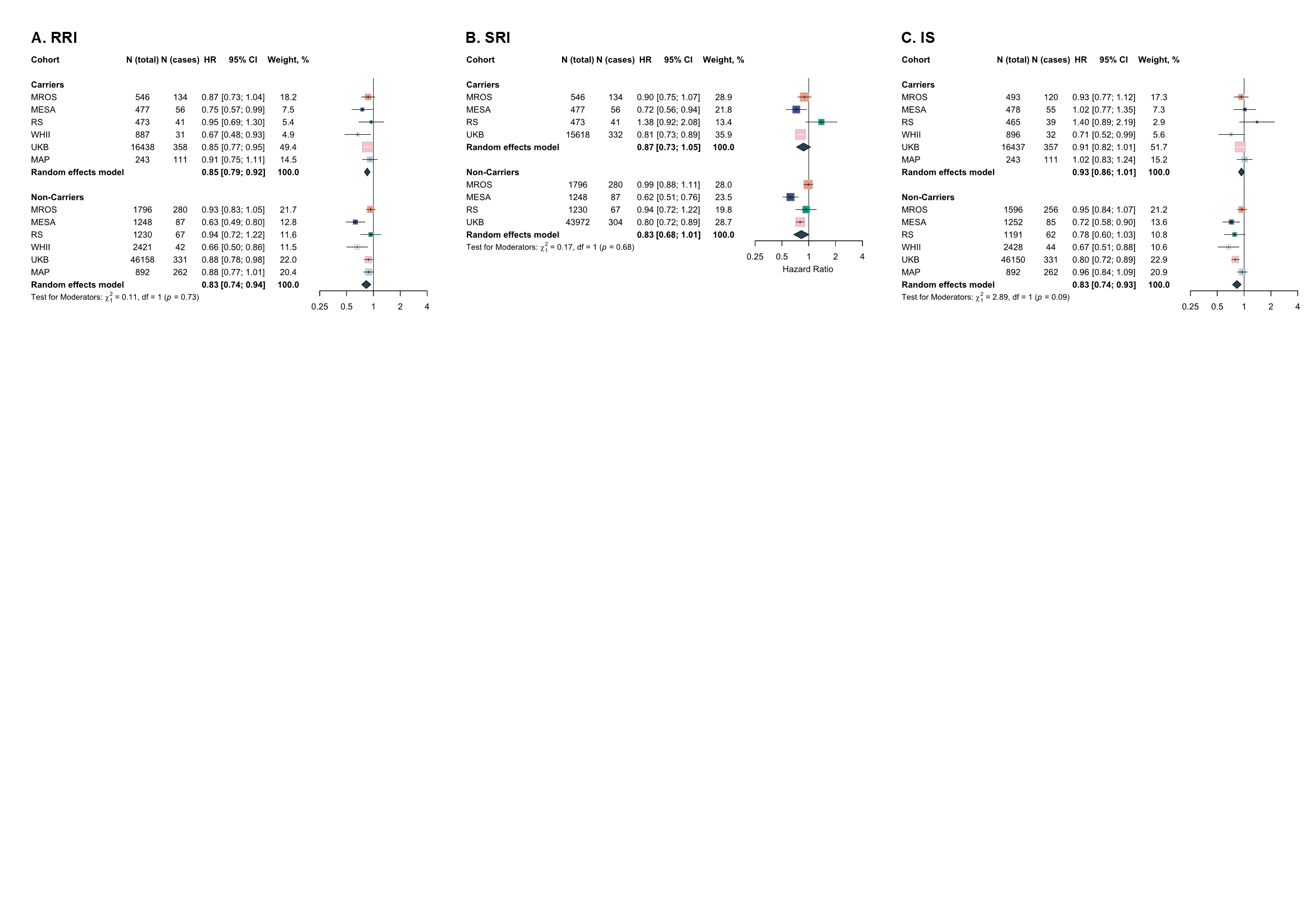
Supplementary Figure 6: Association of 24-hour sleep-wake regularity with risk of all-cause dementia stratified by APOE4-carriership

**Note:** Effect sizes reflect hazard ratios with 95% confidence intervals per on SD higher regularity score. Estimated with Cox Proportional Hazard models, adjusted for age, sex, race/ethnicity, study site, and education. Study-level estimates were pooled using random effects meta-analyses and meta-regression was conducted to explore the moderating effects of sex. RRI: Rest Regularity Index; SRI: Sleep Regularity Index; IS: Interdaily Stability; MrOS: Osteoporotic Fractures in Men Study; RS: Rotterdam Study; MESA: Multi-Ethnic Study of Atherosclerosis; WH: Whitehall II; UKB: UK Biobank; MAP: Rush Memory and Ageing Project.

### Supplementary Figure 7: Non-linearity: restricted cubic-spline models

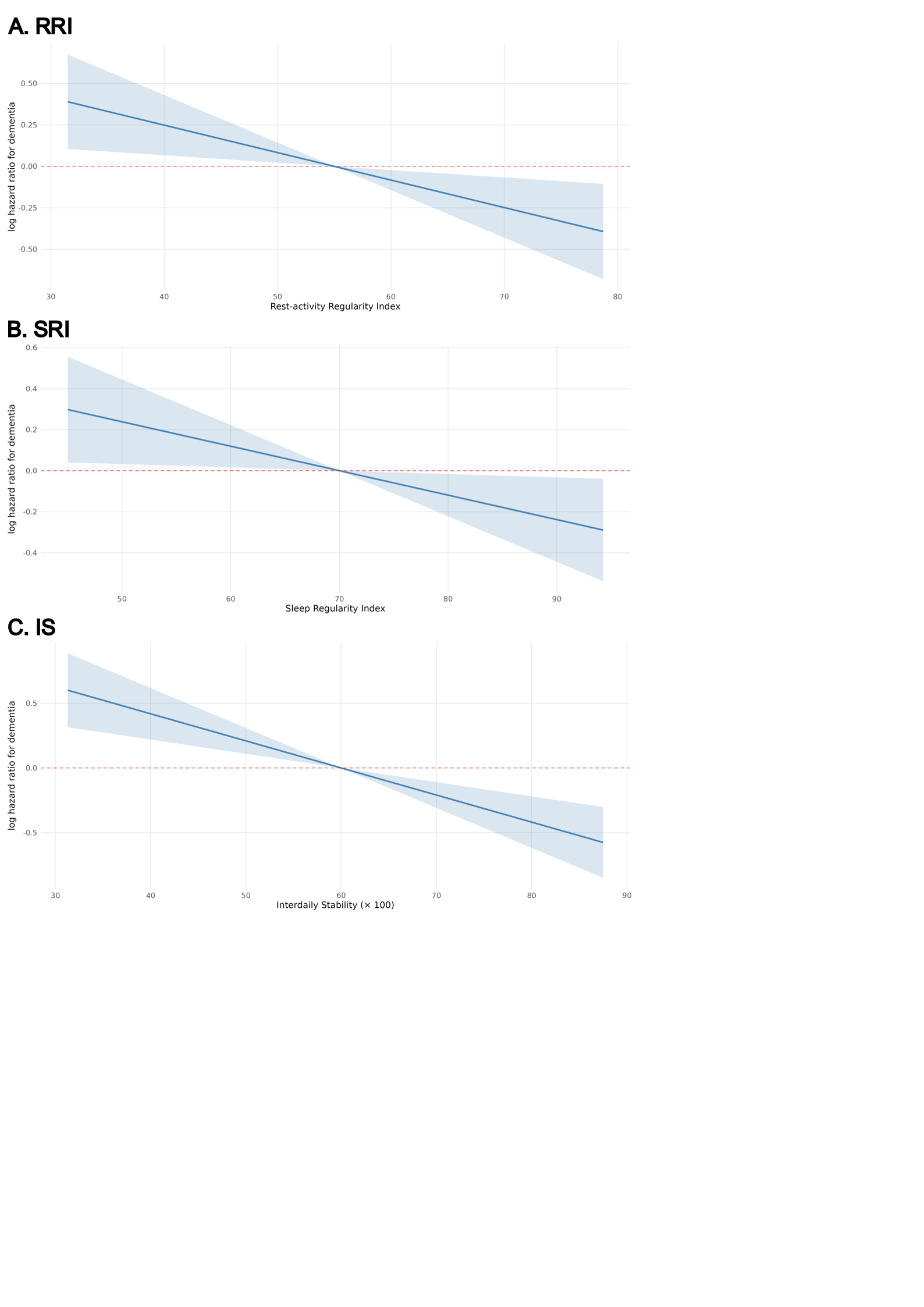

**Note:** Figure displays log hazard ratios with 95% confidence interval for incident dementia; non-linearity was evaluated using restricted cubic splines with knots at the 10^th^, 50^th^, and 90^th^ percentiles, but the association is nevertheless approximated by a straight line. Results are displayed for UKB only as indications for non-linearity were strongest in this cohort. Cox proportional hazard models were adjusted for model 2 covariates.

### Supplementary Figure 8: Fine-Gray competing-risk models (Sensitivity analysis 1)

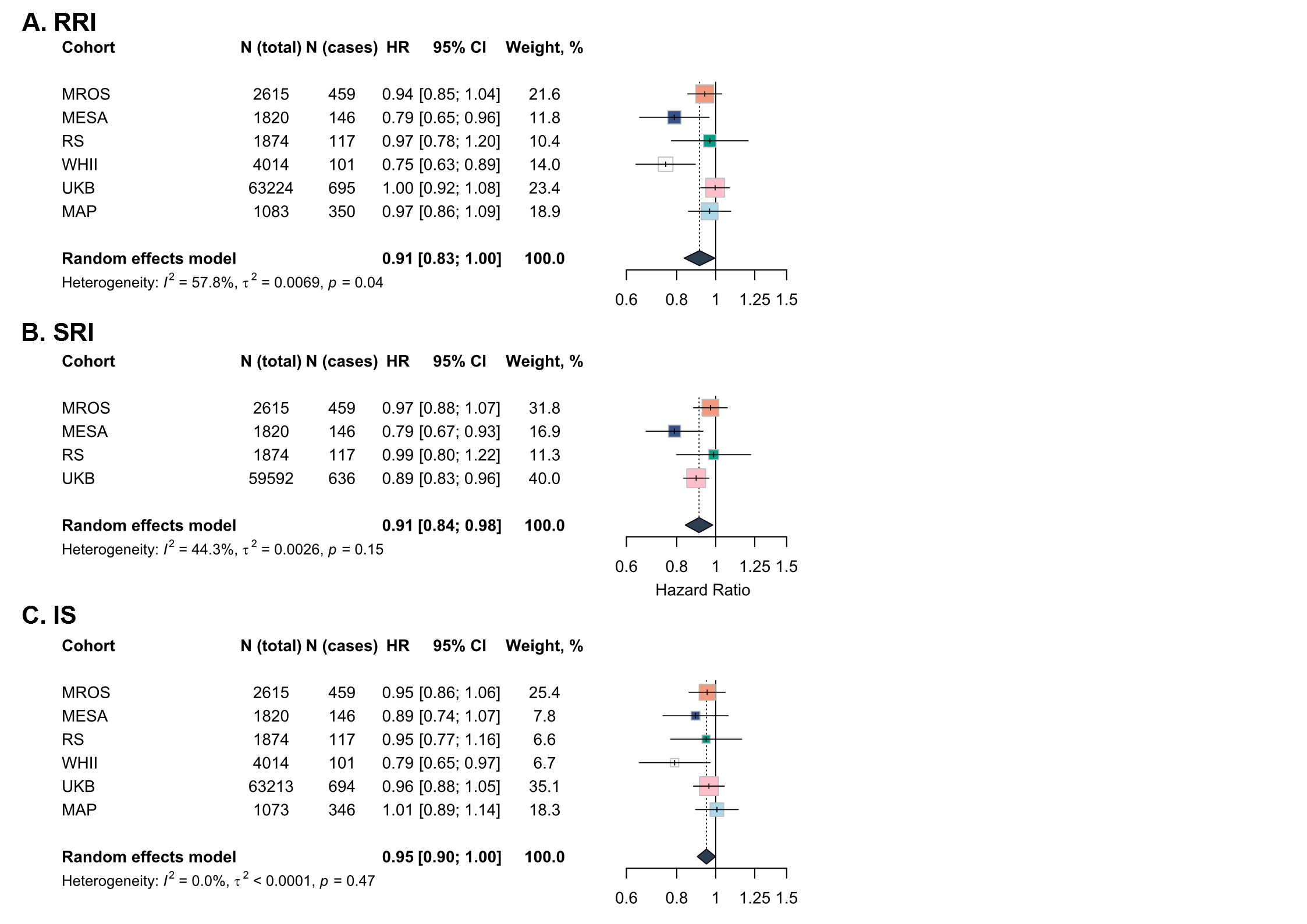

**Note:** Effect sizes reflect hazard ratios with 95% confidence intervals per one SD higher regularity score. Estimated with Fine-Gray models accounting for competing risk. Models were adjusted for age, sex, race/ethnicity, study site, education, marital status, paid employment, body mass index, alcohol use, physical activity, smoking, vascular comorbidities, depressive symptoms, and use of sleep medication. Study-level estimates were pooled using random effects meta-analyses. RRI: Rest Regularity Index; SRI: Sleep Regularity Index; IS: Interdaily Stability; MrOS: Osteoporotic Fractures in Men Study; RS: Rotterdam Study; MESA: Multi-Ethnic Study of Atherosclerosis; WH: Whitehall II; UKB: UK Biobank; MAP: Rush Memory and Ageing Project.

### Supplementary Figure 9: Excluding cohorts relying on hospitalization-based dementia ascertainment (Sensitivity analysis 2)

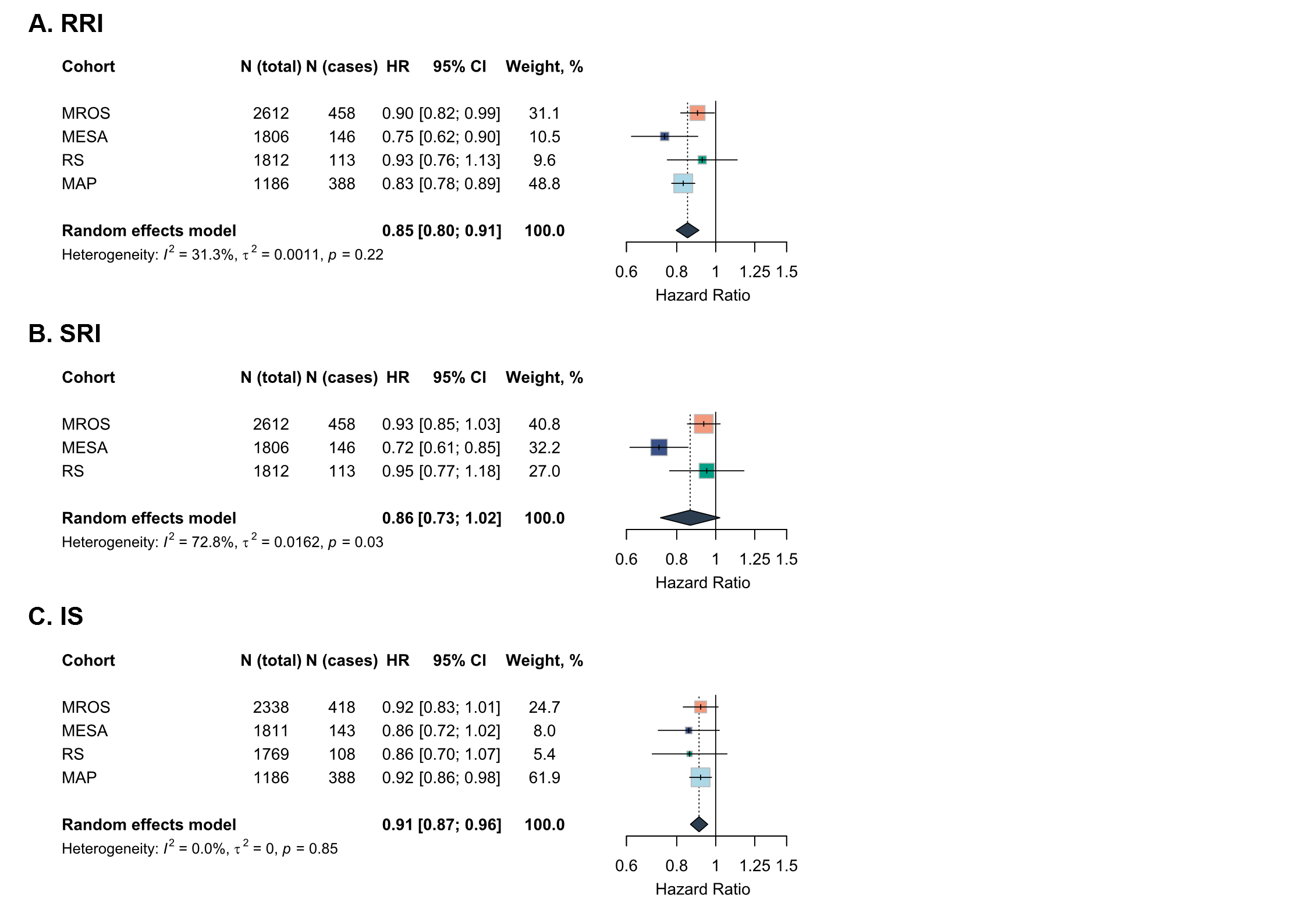

**Note:** Effect sizes reflect hazard ratios with 95% confidence intervals per SD increase in regularity. Estimated with Cox proportional hazard models. Models were adjusted for age, sex, race/ethnicity, study site, education, marital status, paid employment, body mass index, alcohol use, physical activity, smoking, vascular comorbidities, depressive symptoms, and use of sleep medication. Study-level estimates were pooled using random effects meta-analyses. RRI: Rest Regularity Index; SRI: Sleep Regularity Index; IS: Interdaily Stability; MrOS: Osteoporotic Fractures in Men Study; RS: Rotterdam Study; MESA: Multi-Ethnic Study of Atherosclerosis; MAP: Rush Memory and Ageing Project.

### Supplementary Figure 10: Adjusted for sleep duration (Sensitivity analysis 3)

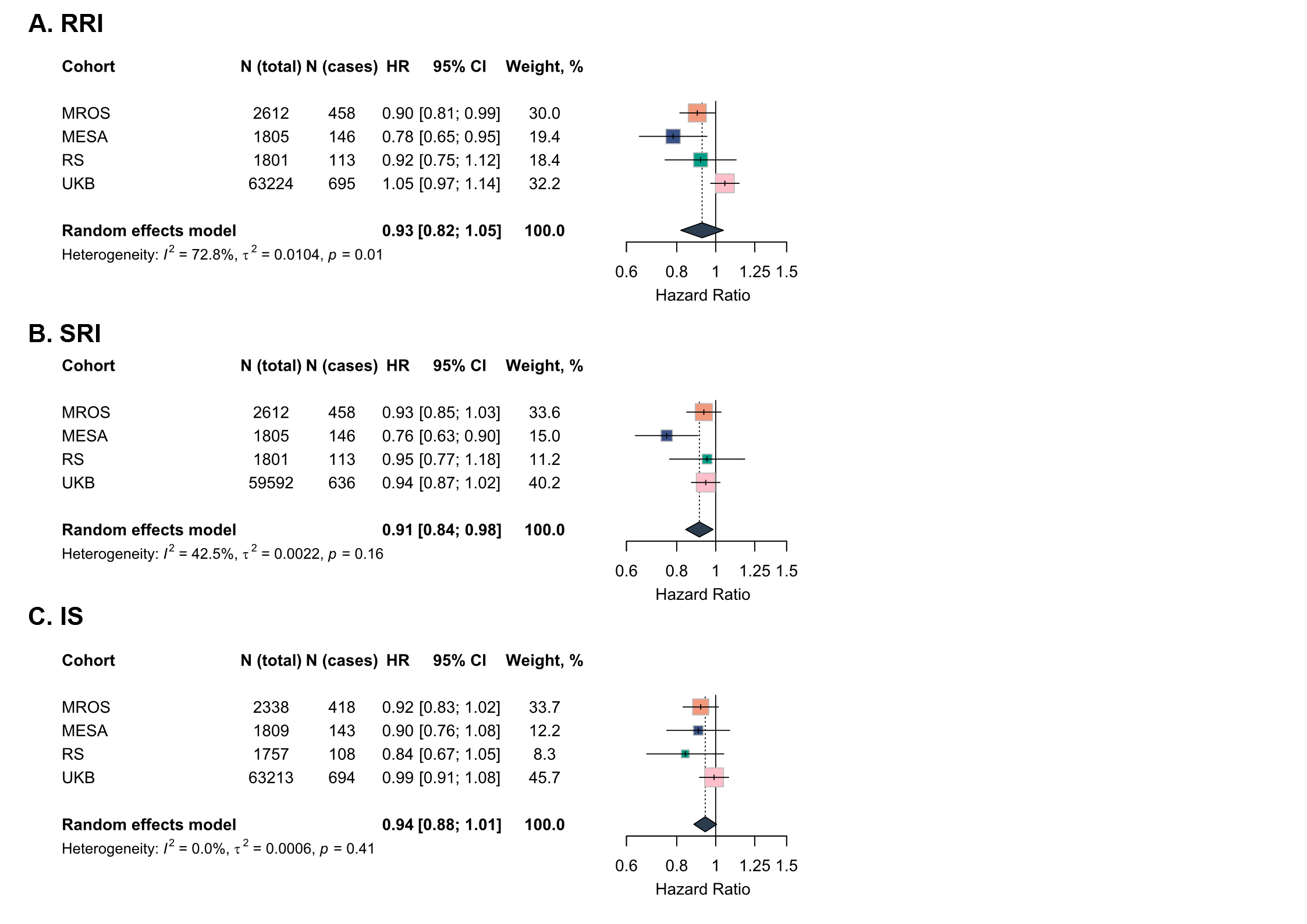

**Note:** Effect sizes reflect hazard ratios with 95% confidence intervals per SD increase in regularity. Estimated with Cox proportional hazard models. Models were adjusted for age, sex, race/ethnicity, study site, education, marital status, paid employment, body mass index, alcohol use, physical activity, smoking, vascular comorbidities, depressive symptoms, use of sleep medication, and sleep duration. Study-level estimates were pooled using random effects meta-analyses. RRI: Rest Regularity Index; SRI: Sleep Regularity Index; IS: Interdaily Stability; MrOS: Osteoporotic Fractures in Men Study; RS: Rotterdam Study; MESA: Multi-Ethnic Study of Atherosclerosis; MAP: Rush Memory and Ageing Project.

### Supplementary Figure 11: Predicted Cognitive Trajectories Across Quartiles of Regularity Metrics in MrOS

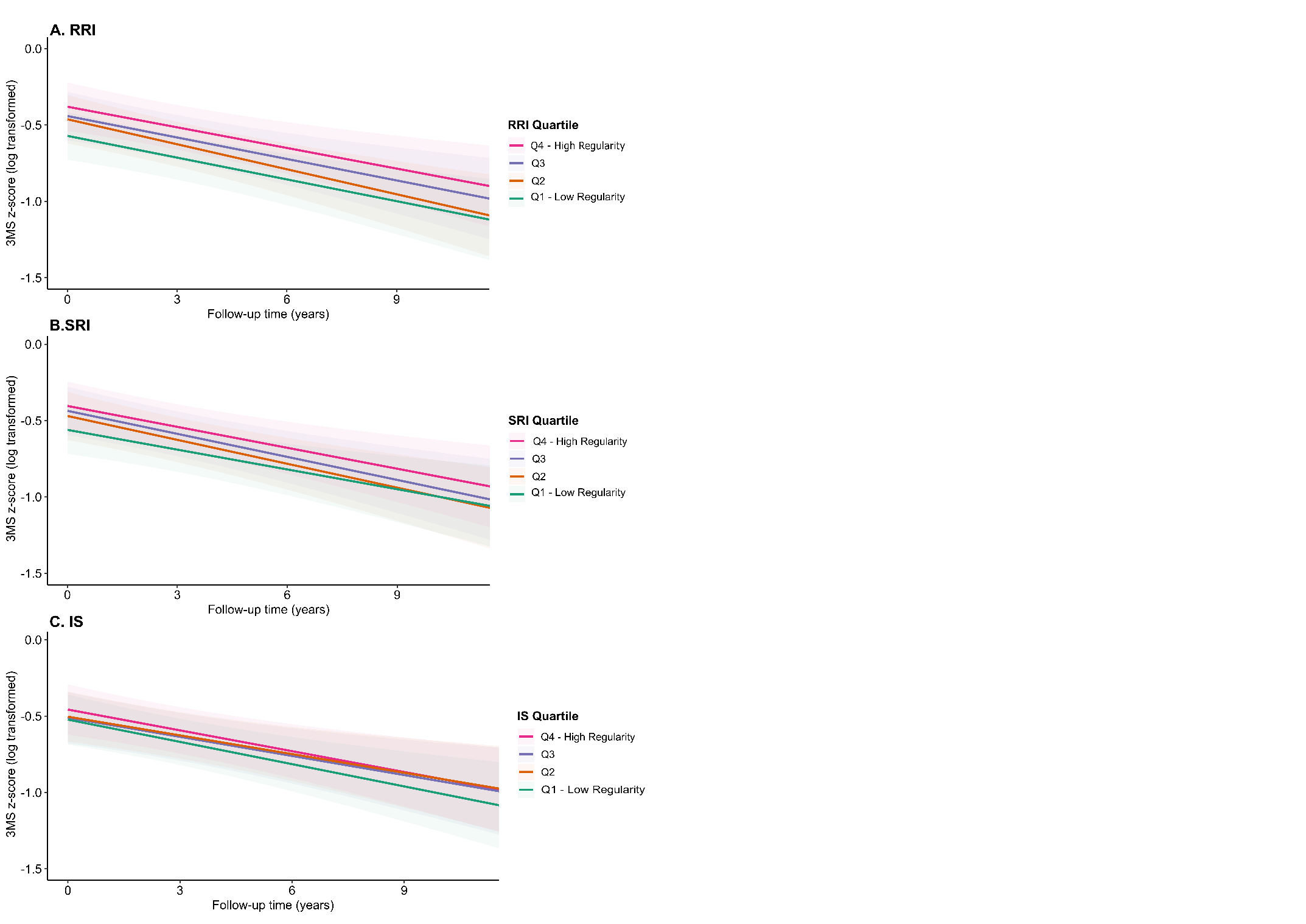

**Note:** Panels A–C present model‑based predicted trajectories of global cognitive performance (3MSS z‑scores) for the MROS cohort, estimated using linear mixed‑effects models. Panel A shows trajectories across quartiles of the Relative Regularity Index (RRI), Panel B across quartiles of the Sleep Regularity Index (SRI), and Panel C across quartiles of the Interdaily Stability (IS) metric. Lines represent predicted values for each quartile (Q1–Q4), and shaded bands indicate 95% confidence intervals. All predictions are adjusted for age, sex, race/ethnicity, study site, education, marital status, paid employment, body mass index, alcohol use, physical activity, smoking, vascular comorbidities, depressive symptoms, and use of sleep medication.

### Supplementary Figure 12: Predicted Cognitive Trajectories Across Quartiles of Regularity Metrics in RS

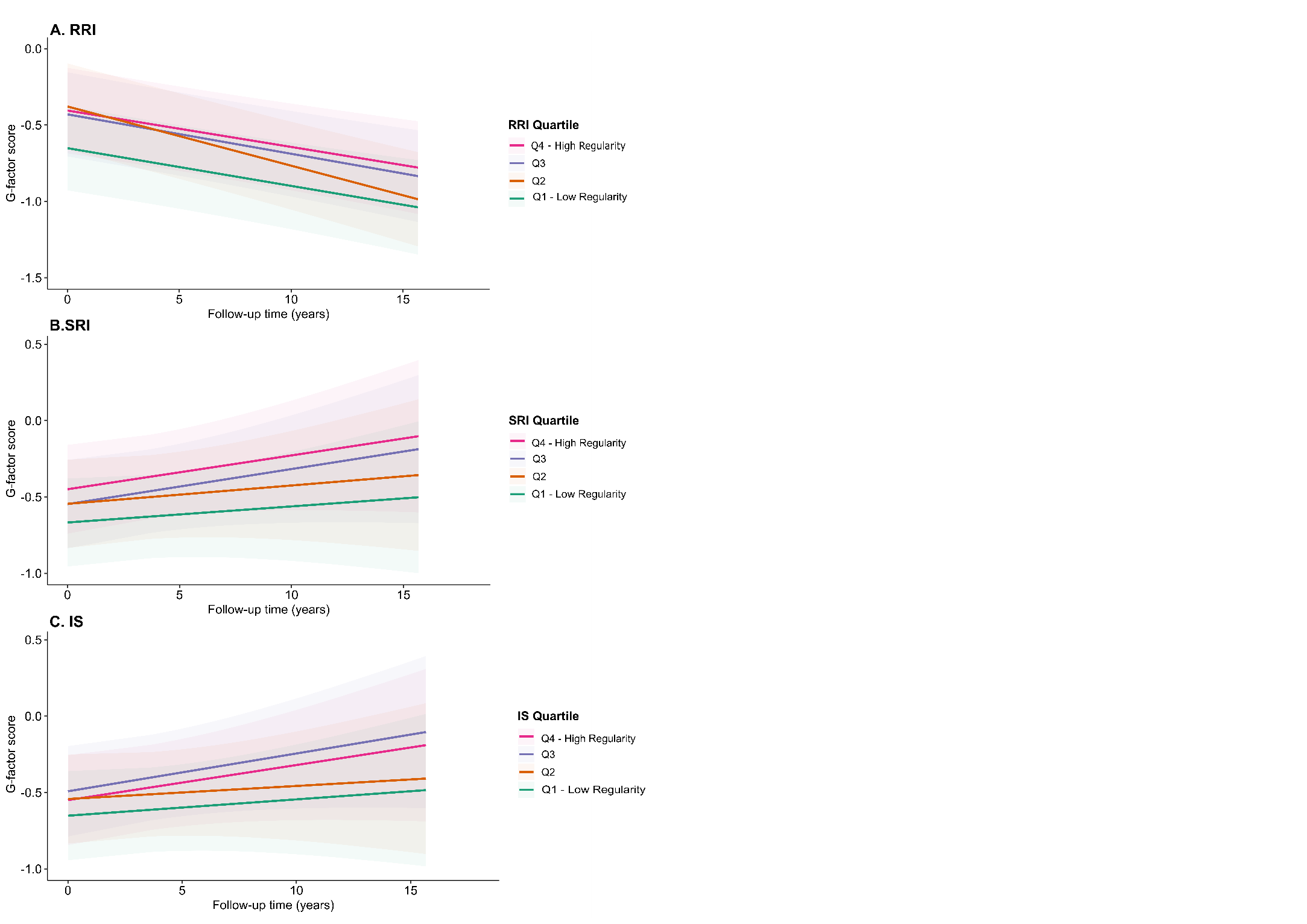

**Note:** Panels A–C present model‑based predicted trajectories of global cognitive performance (G-factor) for the RS cohort, estimated using linear mixed‑effects models. Panel A shows trajectories across quartiles of the Relative Regularity Index (RRI), Panel B across quartiles of the Sleep Regularity Index (SRI), and Panel C across quartiles of the Interdaily Stability (IS) metric. Lines represent predicted values for each quartile (Q1–Q4), and shaded bands indicate 95% confidence intervals. All predictions are adjusted for age, sex, race/ethnicity, study site, education, marital status, paid employment, body mass index, alcohol use, physical activity, smoking, vascular comorbidities, depressive symptoms, and use of sleep medication.

### Supplementary Figure 13: Predicted Cognitive Trajectories Across Quartiles of Regularity Metrics in MAP

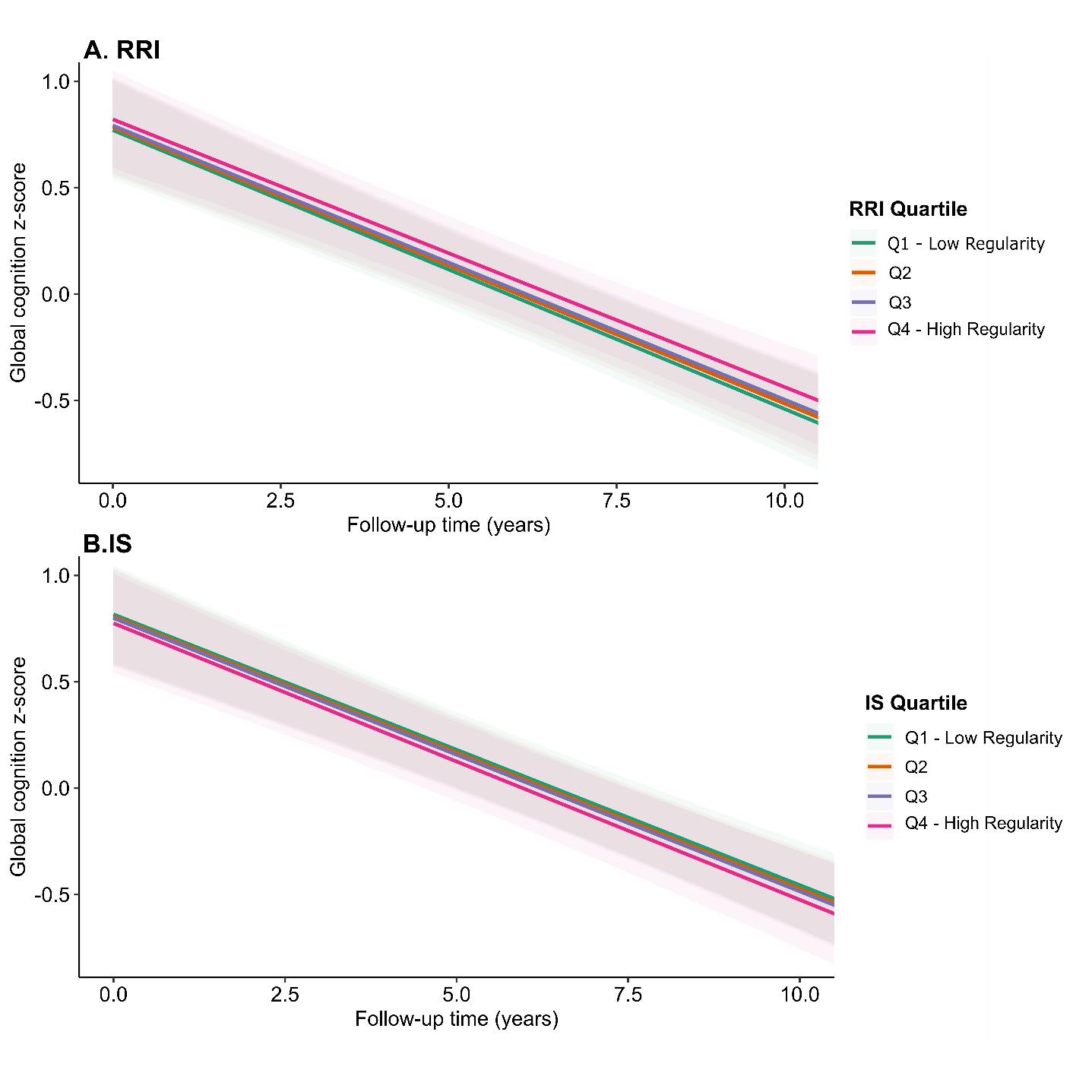

**Note:** Panels A–B present model‑based predicted trajectories of global cognitive performance for the MAP cohort, estimated using linear mixed‑effects models. Panel A shows trajectories across quartiles of the Relative Regularity Index (RRI), Panel B across quartiles of the Interdaily Stability (IS) metric. Lines represent predicted values for each quartile (Q1–Q4), and shaded bands indicate 95% confidence intervals. All predictions are adjusted for age, sex, race/ethnicity, study site, education, marital status, paid employment, body mass index, alcohol use, physical activity, smoking, vascular comorbidities, depressive symptoms, and use of sleep medication.

### Supplementary Table 1: Regularity metrics across cohorts and within subgroups

|  | **MrOS** | **RS** | **MESA** | **MAP** | **Whitehall II** | **UK Biobank** |
| --- | --- | --- | --- | --- | --- | --- |
| RRI | N = 2,612 | N = 1,812 | N = 1,806 | N = 1,186 | N = 3,992 | N = 63,224 |
| *Full sample* | 65.3 [55.2, 73.7] | 57.9 [51.8, 62.0] | 50.3 [39.0, 60.0] | 56.4 [47.9, 62.7] | 61.1 [53.4, 68.0] | 61.3 [54.1, 67.4] |
| *Males* | 65.3 [55.2, 73.7] | 58.7 [53.1, 63.2] | 47.7 [36.6, 57.9] | 57.6 [49.2, 63.8] | 60.4 [52.7, 67.3] | 58.8 [51.4, 65.1] |
| *Females* | n.a. | 57.2 [51.2, 61.3] | 52.0 [41.4, 61.6] | 56.2 [47.5, 62.1] | 63.1 [55.5, 70.1] | 63.2 [56.6, 68.9] |
| *< 65 years* | n.a. | 58.4 [53.0, 62.5] | 55.2 [45.0, 64.2] | 56.0 [50.8, 61.8] | 62.1 [54.6, 68.7] | 61.6 [54.6, 67.6] |
| *65-75 years* | 66.5 [57.7, 74.9] | 57.4 [51.3, 61.8] | 50.1 [38.6, 60.5] | 58.4 [49.2, 64.1] | 60.8 [53.4, 68.1] | 61.1 [53.8, 67.3] |
| *75-85 years* | 64.2 [53.8, 72.6] | 54.7 [48.6, 60.0] | 47.5 [37.9, 57.0] | 56.8 [49.6, 62.8] | 60.5 [52.4, 67.1] | 60.7 [52.4, 67.1] |
| *85+* | 60.8 [50.7, 71.9] | 49.7 [47.1, 54.5] | 41.9 [34.0, 50.4] | 54.5 [42.6, 60.9] | n.a. | n.a. |
| SRI | N = 2,612 | N = 1,812 | N = 1,806 |  |  | N = 59,592 |
| *Full sample* | 82.9 [74.3, 88.5] | 84.3 [78.6, 88.8] | 70.5 [58.2, 78.9] | n.a. | n.a. | 80.6 [73.1, 86.1] |
| *Males* | 82.9 [74.3, 88.5] | 84.3 [78.6, 89.0] | 68.4 [56.8, 78.2] |  |  | 80.0 [72.3, 85.8] |
| *Females* | n.a. | 84.3 [78.5, 88.8] | 71.9 [59.5, 79.4] |  |  | 81.0 [73.8, 86.3] |
| *< 65 years* | n.a. | 83.4 [77.4, 87.8] | 72.5 [62.1, 79.8] |  |  | 80.9 [73.6, 86.1] |
| *65-75 years* | 83.1 [75.5, 88.6] | 86.1 [81.3, 91.0] | 70.5 [58.2, 79.1] |  |  | 80.3 [72.8, 86.1] |
| *75-85 years* | 82.6 [73.8, 88.4] | 86.6 [81.6, 90.2] | 68.0 [57.0, 78.2] |  |  | 79.2 [71.1, 85.8] |
| *85+* | 83.4 [71.8, 88.5] | 84.1 [76.9, 86.8] | 61.2 [47.7, 74.1] |  |  | n.a. |
| *IS* | N = 2,338 | N = 1,769 | N = 1,811 | N = 1,170 | N = 4,014 | N = 63,213 |
| *Full sample* | 0.75 [0.69, 0.81] | 0.80 [0.72, 0.85] | 0.57 [0.49, 0.65] | 0.53 [0.44; 0.62] | 0.54 [0.45, 0.61] | 0.68 [0.60, 0.75] |
| *Males* | 0.75 [0.69, 0.81] | 0.79 [0.71, 0.85] | 0.56 [0.48, 0.63] | 0.51 [0.41, 0.60] | 0.53 [0.44, 0.60] | 0.66 [0.58, 0.74] |
| *Females* | n.a. | 0.80 [0.73, 0.86] | 0.59 [0.51, 0.66] | 0.54 [0.45, 0.62] | 0.55 [0.46, 0.62] | 0.70 [0.62, 0.76] |
| *< 65 years* | n.a. | 0.79 [0.71, 0.85] | 0.57 [0.49, 0.65] | 0.51 [0.40, 0.57] | 0.52 [0.44, 0.59] | 0.67 [0.59, 0.74] |
| *65-75 years* | 0.76 [0.69, 0.81] | 0.81 [0.74, 0.87] | 0.58 [0.50, 0.65] | 0.52 [0.43, 0.59] | 0.54 [0.46, 0.61] | 0.69 [0.62, 0.76] |
| *75-85 years* | 0.75 [0.68, 0.81] | 0.80 [0.73, 0.86] | 0.58 [0.49, 0.65] | 0.54 [0.44, 0.61] | 0.53 [0.44, 0.61] | 0.69 [0.61, 0.75] |
| *85+* | 0.75 [0.68, 0.81] | 0.80 [0.71, 0.86] | 0.57 [0.46, 0.64] | 0.54 [0.45, 0.63] | n.a. | n.a. |

**Note:** Median [Q1, Q3]**;** RRI: Rest Regularity Index; SRI: Sleep Regularity Index; IS: Interdaily Stability MrOS: Osteoporotic Fractures in Men Study; RS: Rotterdam Study; MESA: Multi-Ethnic Study of Atherosclerosis; MAP: Rush Memory and Aging Project; WH: Whitehall II; UKB: UK Biobank;

### Supplementary Table 2: Correlations between regularity indices

|  |  | Spearman Correlation | | |
| --- | --- | --- | --- | --- |
|  |  | IS | RRI | SRI |
| MrOS | IS | 1.00 | 0.58 | 0.56 |
|  | RRI | - | 1.00 | 0.77 |
|  | SRI | - | - | 1.00 |
| RS | IS | 1.00 | 0.44 | 0.62 |
|  | RRI | - | 1.00 | 0.44 |
|  | SRI | - | - | 1.00 |
| MESA | IS | 1.00 | 0.65 | 0.57 |
|  | RRI | - | 1.00 | 0.75 |
|  | SRI | - | - | 1.00 |
| MAP | IS | 1.00 | 0.44 | - |
|  | RRI | - | 1.00 | - |
|  | SRI | - | - | - |
| Whitehall II | IS | 1.00 | 0.35 | - |
|  | RRI | - | 1.00 | - |
|  | SRI | - | - | - |
| UK Biobank | IS | 1.00 | 0.49 | 0.40 |
|  | RRI | - | 1.00 | 0.72 |
|  | SRI | - | - | 1.00 |

**Note:** Correlations between regularity indices were estimated with Spearman correlations. RRI: Rest Regularity Index; SRI: Sleep Regularity Index; IS: Interdaily Stability MrOS: Osteoporotic Fractures in Men Study; RS: Rotterdam Study; MESA: Multi-Ethnic Study of Atherosclerosis; MAP: Rush Memory and Aging Project.

### Supplementary Table 3. Quartile Transition Matrices

A. **RRI x SRI**

|  |  |  | SRI |  |  |  |  |
| --- | --- | --- | --- | --- | --- | --- | --- |
|  |  |  | Q4 | Q3 | Q2 | Q1 | **No transition^a^** |
| MrOS | RRI | Q4 | **419 (64.2%)** | 201 (30.8%) | 32 (4.9%) | 1 (0.2%) | **1389 (53.2%)** |
|  |  | Q3 | 164 (25.2%) | **253 (38.8%)** | 200 (30.7%) | 35 (5.4%) |  |
|  |  | Q2 | 57 (8.7%) | 156 (23.9%) | **270 (41.3%)** | 171 (26.1%) |  |
|  |  | Q1 | 10 (1.5%) | 43 (6.6%) | 153 (23.4%) | **447 (68.5%)** |  |
| RS | RRI | Q4 | **195 (43%)** | 130 (28.7%) | 88 (19.4%) | 40 (8.8%) | **681 (37.6%)** |
|  |  | Q3 | 140 (30.9%) | **130 (28.7%)** | 105 (23.2%) | 78 (17.2%) |  |
|  |  | Q2 | 82 (18.1%) | 118 (26%) | **137 (30.2%)** | 116 (25.6%) |  |
|  |  | Q1 | 36 (7.9%) | 75 (16.6%) | 123 (27.2%) | **219 (48.3%)** |  |
| MESA | RRI | Q4 | **281 (62.3%)** | 131 (29.0%) | 37 (8.2%) | 2 (0.4%) | **938 (51.9%)** |
|  |  | Q3 | 115 (25.4%) | **174 (38.5%)** | 131 (29.0%) | 32 (7.1%) |  |
|  |  | Q2 | 46 (10.2%) | 118 (26.2%) | **176 (39.0%)** | 111 (24.6%) |  |
|  |  | Q1 | 10 (2.2%) | 28 (6.2%) | 107 (23.7%) | **307 (67.9%)** |  |
| UK Biobank | RRI | Q4 | **9372 (59.7%)** | 4701 (29.9%) | 1462 (9.3%) | 165 (1.1%) | **30946 (50.2%)** |
|  |  | Q3 | 4045 (26%) | **5703 (36.7%)** | 4634 (29.8%) | 1160 (7.5%) |  |
|  |  | Q2 | 1646 (10.7%) | 3819 (24.9%) | **5831 (38%)** | 4059 (26.4%) |  |
|  |  | Q1 | 350 (2.3%) | 1211 (8%) | 3497 (23.2%) | **10040 (66.5%)** |  |

B. **RRI x IS**

|  |  |  | IS |  |  |  |  |
| --- | --- | --- | --- | --- | --- | --- | --- |
|  |  |  | Q4 | Q3 | Q2 | Q1 | **No transition^a^** |
| MrOS | RRI | Q4 | **298 (50.3%)** | 176 (29.7%) | 86 (14.5%) | 32 (5.4%) | **1038 (44.5%)** |
|  |  | Q3 | 164 (27.6%) | **199 (33.4%)** | 158 (26.6%) | 74 (12.4%) |  |
|  |  | Q2 | 93 (15.8%) | 152 (25.8%) | **203 (34.5%)** | 141 (23.9%) |  |
|  |  | Q1 | 29 (5.2%) | 55 (9.8%) | 137 (24.5%) | **338 (60.5%)** |  |
| RS | RRI | Q4 | **195 (45.7%)** | 101 (23.7%) | 75 (17.6%) | 56 (13.1%) | **680 (39.8%)** |
|  |  | Q3 | 137 (32.2%) | **120 (28.2%)** | 99 (23.3%) | 69 (16.2%) |  |
|  |  | Q2 | 70 (16.4%) | 106 (24.8%) | **150 (35%)** | 102 (23.8%) |  |
|  |  | Q1 | 24 (5.6%) | 58 (13.6%) | 130 (30.4%) | **215 (50.4%)** |  |
| MESA | RRI | Q4 | **255 (56.8%)** | 113 (25.2%) | 57 (12.7%) | 24 (5.3%) | **843 (46.9%)** |
|  |  | Q3 | 133 (29.5%) | **161 (35.7%)** | 115 (25.5%) | 42 (9.3%) |  |
|  |  | Q2 | 51 (11.4%) | 125 (27.9%) | **158 (35.3%)** | 114 (25.4%) |  |
|  |  | Q1 | 8 (1.8%) | 50 (11.1%) | 122 (27.2%) | **269 (59.9%)** |  |
| MAP | RRI | Q4 | **150 (43.6%)** | 99 (28.8%) | 58 (16.9%) | 37 (10.8%) | **542 (39.4%)** |
|  |  | Q3 | 101 (29.4%) | **106 (30.8%)** | 90 (26.2%) | 47 (13.7%) |  |
|  |  | Q2 | 69 (20.1%) | 94 (27.3%) | **104 (30.2%)** | 77 (22.4%) |  |
|  |  | Q1 | 26 (7.6%) | 44 (12.8%) | 92 (26.7%) | **182 (52.9%)** |  |
| Whitehall II | RRI | Q4 | **408 (40.8%)** | 268 (26.8%) | 198 (19.8%) | 125 (12.5%) | **1390 (34.8%)** |
|  |  | Q3 | 279 (27.9%) | **280 (28.0%)** | 263 (26.3%) | 177 (17.7%) |  |
|  |  | Q2 | 195 (19.5%) | 265 (26.5%) | **272 (27.2%)** | 267 (26.7%) |  |
|  |  | Q1 | 120 (12.0%) | 174 (17.4%) | 276 (27.6%) | **430 (43.0%)** |  |
| UK Biobank | RRI | Q4 | **7686 (47.0%)** | 4577 (28.0%) | 2839 (17.3%) | 1264 (7.7%) | **25879 (39.5%)** |
|  |  | Q3 | 4695 (28.7%) | **4836 (29.5%)** | 4257 (26.0%) | 2580 (15.8%) |  |
|  |  | Q2 | 2876 (17.6%) | 4369 (26.7%) | **4962 (30.3%)** | 4160 (25.4%) |  |
|  |  | Q1 | 1072 (6.5%) | 2456 (15.0%) | 4445 (27.2%) | **8395 (51.3%)** |  |

C. **SRI x IS**

|  |  |  | IS |  |  |  |  |
| --- | --- | --- | --- | --- | --- | --- | --- |
|  |  |  | Q4 | Q3 | Q2 | Q1 | **No transition^a^** |
| MrOS | SRI | Q4 | **299 (49.4%)** | 169 (27.9%) | 96 (15.9%) | 41 (6.8%) | **1002 (42.9%)** |
|  |  | Q3 | 169 (28.8%) | **196 (33.4%)** | 150 (25.6%) | 71 (12.1%) |  |
|  |  | Q2 | 85 (14.5%) | 147 (25.1%) | **194 (33.1%)** | 160 (27.3%) |  |
|  |  | Q1 | 31 (5.6%) | 70 (12.5%) | 144 (25.8%) | **313 (56.1%)** |  |
| RS | SRI | Q4 | **250 (57.2%)** | 115 (26.3%) | 53 (12.1%) | 19 (4.3%) | **793 (46.5%)** |
|  |  | Q3 | 117 (26.9%) | **133 (30.6%)** | 111 (25.5%) | 74 (17%) |  |
|  |  | Q2 | 49 (11.6%) | 100 (23.7%) | **167 (39.6%)** | 106 (25.1%) |  |
|  |  | Q1 | 10 (2.4%) | 37 (9%) | 123 (29.8%) | **243 (58.8%)** |  |
| MESA | SRI | Q4 | **234 (52.0%)** | 121 (26.9%) | 74 (16.4%) | 21 (4.7%) | **878 (42.3%)** |
|  |  | Q3 | 123 (27.3%) | **146 (32.4%)** | 121 (26.8%) | 61 (13.5%) |  |
|  |  | Q2 | 66 (14.7%) | 128 (28.5%) | **134 (29.8%)** | 121 (26.9%) |  |
|  |  | Q1 | 24 (5.4%) | 54 (12.1%) | 123 (27.5%) | **246 (55.0%)** |  |
| UK Biobank | SRI | Q4 | **6576 (42.7%)** | 4345 (28.2%) | 3021 (19.6%) | 1468 (9.5%) | **22566 (36.6%)** |
|  |  | Q3 | 4353 (28.2%) | **4431 (28.7%)** | 4033 (26.1%) | 2613 (16.9%) |  |
|  |  | Q2 | 3072 (19.9%) | 3991 (25.9%) | **4517 (29.3%)** | 3843 (24.9%) |  |
|  |  | Q1 | 1670 (10.8%) | 2662 (17.3%) | 4048 (26.2%) | **7042 (45.7%)** |  |

**Note:** Numbers reflect N (%) for each combination of quartiles for each combination of regularity indices. ^a^participants were scored in the same quartile for both indices. RRI: Rest Regularity Index; SRI: Sleep Regularity Index; IS: Interdaily Stability MrOS: Osteoporotic Fractures in Men Study; RS: Rotterdam Study; MESA: Multi-Ethnic Study of Atherosclerosis; MAP: Rush Memory and Aging Project.

### Supplementary Table 4: Association of rhythm regularity with risk of dementia

|  | **MrOS** | **RS** | **MESA** | **MAP** | **Whitehall II** | **UK Biobank** | **Pooled HR** |
| --- | --- | --- | --- | --- | --- | --- | --- |
| Number of dementia cases *N (%)* | 459 (17.6%) | 117 (6.2%) | 146 (8.0%) | 388 (32.7%) | 101 (2.5%) | 695 (1.1%) | **1,906** |
| Follow-up, *mean (SD)* |  |  |  |  |  |  |  |
| *Full sample* | 7.1 (3.8) | 11.6 (3.2) | 9.0 (2.7) | 7.2 (4.2) | 6.9 (1.0) | 7.9 (1.1) |  |
| *< 65 years* | - | 12.2 (2.4) | 10.2 (1.9) | - | 7.08 (0.7) | 8.1 (0.9) |  |
| *65-75 years* | 8.40 (3.5) | 11.9 (3.5) | 9.2 (2.6) | 9.14 (5.2) | 6.97 (0.9) | 7.8 (1.2) |  |
| *75-85 years* | 6.34 (3.8) | 8.16 (4.3) | 8.1 (2.8) | 7.92 (4.3) | 6.58 (1.5) | 7.3 (1.4) |  |
| *85+* | 4.08 (2.8) | 5.04 (3.6) | 6.2 (2.9) | 5.71 (3.3) | - | - |  |
| **RRI** |  |  |  |  |  |  |  |
|  | HR (95%CI) | | | | | | |
| **Full Sample** |  |  |  |  |  |  |  |
| *Model 1* | 0.90 (0.82–0.99) | 0.91 (0.74–1.10) | 0.70 (0.59–0.85) | 0.86 (0.77–0.95) | 0.71 (0.60-0.85) | 0.87 (0.81–0.94) | **0.84 (0.78-0.90)** |
| *Model 2* | 0.90 (0.82–0.99) | 0.93 (0.76–1.13) | 0.75 (0.62–0.90) | 0.83 (0.78–0.89) | 0.74 (0.61-0.89) | 0.99 (0.91–1.07) | **0.86 (0.79-0.95)** |
| **Sex** |  |  |  |  |  |  |  |
| *Males* | 0.90 (0.82–0.99) | 0.81 (0.60–1.08) | 0.54 (0.41–0.71) | 0.81 (0.64–1.01) | 0.77 (0.61–0.96) | 0.87 (0.79–0.96) | **0.79 (0.70-0.91)** |
| *Females* | - | 1.01 (0.77–1.32) | 0.91 (0.71–1.17) | 0.88 (0.78–0.98) | 0.62 (0.46–0.83) | 0.88 (0.78–0.99) | **0.87 (0.81-0.94)** |
| **Age** |  |  |  |  |  |  |  |
| *< 65 years* | - | 0.91 (0.57–1.44) | 0.42 (0.24–0.75) | - | - | 0.72 (0.59–0.89) | **0.69 (0.49-0.96)** |
| *65-75 years* | 0.80 (0.68–0.94) | 0.71 (0.55–0.92) | 0.69 (0.49–0.97) | 1.08 (0.73–1.60) | 0.76 (0.57-1.01) | 0.90 (0.83–0.97) | **0.83 (0.75-0.92)** |
| *75-85 years* | 0.96 (0.85–1.08) | 1.23 (0.85–1.79) | 0.64 (0.48–0.85) | 0.90 (0.77–1.05) | 0.72 (0.57-0.92) | 0.84 (0.64–1.11) | **0.86 (0.74-0.99)** |
| *85+* | 0.95 (0.71–1.26) | 0.62 (0.23–1.69) | 1.13 (0.70–1.82) | 0.78 (0.67–0.91) | - | - | 0.85 (0.71-1.02) |
| **APOE4-carriership** |  |  |  |  |  |  |  |
| *APOE4-carrier* | 0.87 (0.73–1.04) | 0.95 (0.69–1.30) | 0.75 (0.57–0.99) | 0.89 (0.76–1.05) | 0.67 (0.48–0.93) | 0.85 (0.77–0.95) | **0.85 (0.79-0.92)** |
| *APOE4-non carrier* | 0.93 (0.83–1.05) | 0.94 (0.72–1.22) | 0.63 (0.49–0.80) | 0.83 (0.76–0.89) | 0.66 (0.50–0.86) | 0.88 (0.78–0.98) | **0.83 (0.74-0.94)** |
| **SRI** |  |  |  |  |  |  |  |
| **Full Sample** |  |  |  |  |  |  |  |
| *Model 1* | 0.93 (0.85–1.02) | 0.99 (0.80–1.23) | 0.68 (0.58–0.80) | - | - | 0.81 (0.75–0.86) | **0.85 (0.74-0.97)** |
| *Model 2* | 0.93 (0.85–1.03) | 0.95 (0.77–1.18) | 0.72 (0.61–0.85) | - | - | 0.88 (0.82–0.95) | **0.87 (0.79-0.97)** |
| **Sex** |  |  |  |  |  |  |  |
| *Males* | 0.93 (0.85–1.02) | 0.99 (0.71–1.37) | 0.58 (0.47–0.71) | - | - | 0.78 (0.71–0.86) | **0.80 (0.64-1.00)** |
| *Females* | - | 1.03 (0.78–1.38) | 0.88 (0.70–1.12) | - | - | 0.85 (0.76–0.95) | **0.88 (0.79-0.96)** |
| **Age** |  |  |  |  |  |  |  |
| *< 65 years* | n.a. | 1.15 (0.67–1.97) | 0.54 (0.32–0.92) | - | - | 0.70 (0.58–0.84) | 0.74 (0.53-1.02) |
| *65-75 years* | 0.81 (0.69–0.96) | 0.88 (0.65–1.18) | 0.67 (0.50–0.90) | - | - | 0.83 (0.76–0.89) | **0.82 (0.77-0.87)** |
| *75-85 years* | 1.00 (0.88–1.12) | 1.37 (0.89–2.11) | 0.59 (0.46–0.76) | - | - | 0.78 (0.60–1.02) | 0.87 (0.63-1.20) |
| *85+* | 0.96 (0.71–1.31) | 0.69 (0.28–1.71) | 0.99 (0.65–1.49) | - | - | - | 0.95 (0.75-1.20) |
| **APOE4-carriership** |  |  |  |  |  |  |  |
| *APOE4-carrier* | 0.87 (0.73–1.04) | 0.95 (0.69–1.30) | 0.75 (0.57–0.99) | 0.83 (0.76–0.89) | 0.67 (0.48–0.93) | 0.81 (0.73–0.89) | 0.87 (0.73-1.05) |
| *APOE4-non carrier* | 0.93 (0.83–1.05) | 0.94 (0.72–1.22) | 0.63 (0.49–0.80) | 0.89 (0.76–1.05) | 0.66 (0.50–0.86) | 0.80 (0.72–0.89) | 0.83 (0.68-1.01) |
| **IS** |  |  |  |  |  |  |  |
| **Full Sample** |  |  |  |  |  |  |  |
| *Model 1* | 0.92 (0.84–1.01) | 0.87 (0.70–1.07) | 0.85 (0.71–1.00) | 0.96 (0.86–1.06) | 0.76 (0.63-0.92) | 0.85 (0.79–0.91) | **0.88 (0.83-0.93)** |
| *Model 2* | 0.92 (0.83–1.01) | 0.86 (0.69–1.06) | 0.86 (0.72–1.02) | 0.92 (0.86–0.98) | 0.78 (0.65-0.94) | 0.96 (0.88–1.04) | **0.91 (0.88-0.95)** |
| **Sex** |  |  |  |  |  |  |  |
| *Males* | 0.92 (0.84–1.01) | 0.84 (0.62–1.14) | 0.71 (0.56–0.91) | 0.95 (0.78–1.17) | 0.78 (0.62–0.98) | 0.84 (0.76–0.93) | **0.89 (0.84-0.95)** |
| *Females* | - | 0.89 (0.66–1.20) | 1.01 (0.79–1.28) | 0.95 (0.84–1.08) | 0.74 (0.54–1.02) | 0.82 (0.73–0.92) | **0.83 (0.73-0.94)** |
| **Age** |  |  |  |  |  |  |  |
| *< 65 years* | - | 1.01 (0.63–1.63) | 0.97 (0.50–1.85) | - | - | 0.74 (0.61–0.90) | 0.81 (0.65-1.02) |
| *65-75 years* | 0.89 (0.74–1.06) | 0.88 (0.65–1.18) | 0.80 (0.55–1.15) | 1.34 (0.95–1.90) | 0.74 (0.55-1.00) | 0.85 (0.78–0.92) | **0.86 (0.80-0.92)** |
| *75-85 years* | 0.94 (0.83–1.07) | 1.03 (0.62–1.69) | 0.72 (0.56–0.94) | 0.88 (0.76–1.02) | 0.80 (0.63-1.03) | 1.11 (0.81–1.51) | **0.90 (0.83-0.97)** |
| *85+* | 1.02 (0.76–1.37) | 0.16 (0.04–0.70) | 1.03 (0.68–1.55) | 0.93 (0.79–1.10) | - | - | 0.94 (0.82-1.08) |
| **APOE4-carriership** |  |  |  |  |  |  |  |
| *APOE4-carrier* | 0.93 (0.77–1.12) | 1.40 (0.89–2.19) | 1.02 (0.77–1.35) | 0.97 (0.83–1.13) | 0.71 (0.52–0.99) | 0.91 (0.82–1.01) | 0.93 (0.86-1.01) |
| *APOE4-non carrier* | 0.95 (0.84–1.07) | 0.78 (0.60–1.03) | 0.72 (0.58–0.90) | 0.90 (0.84–0.97) | 0.67 (0.51–0.88) | 0.80 (0.72–0.89) | 0.83 (0.74-0.93) |

**Note:** Effect sizes reflect Hazard Ratio (HR) with 95% confidence intervals per SD increase in regularity. Estimated with Cox proportional hazard models. Model 1 was adjusted for age, sex, race/ethnicity, study site and education. Model 2 was additionally adjusted for marital status, paid employment, body mass index, alcohol use, physical activity, smoking, vascular comorbidities, depressive symptoms, and use of sleep medication. Stratified analyses were adjusted for Model 1 covariates. Study-level estimates were pooled using random effects meta-analyses. Statistically significant pooled effect sizes are **bolded.** RRI: Rest Regularity Index; SRI: Sleep Regularity Index; IS: Interdaily Stability MrOS: Osteoporotic Fractures in Men Study; RS: Rotterdam Study; MESA: Multi-Ethnic Study of Atherosclerosis; MAP: Rush Memory and Aging Project;

### Supplementary Table 5: Association of sleep-wake regularity with global cognition over time

|  |  | **MrOS** | **RS** | **MESA** | **MAP** | **Meta-analysis** | |
| --- | --- | --- | --- | --- | --- | --- | --- |
| **Follow-up Length (years)** | | 6.6 (3.8) | 9.9 (6.6) | 6.4 (5.3) | 5.8 (4.0) |  | |
|  | | β (95% CI) | | | | Pooled Effect size | Heterogeneity |
| **RRI** |  |  |  |  |  |  |  |
| **Global Cognition** | Main effects | .053 (.023–.084) | .073 (.030–.117) | -.020 (-.061–.022) | .020 (-.009–.048) | .03 (-.01-.07) | *I^2^*= 75.0%  *p* < 0.01 |
|  | Interaction with follow-up time | .002 (-.002–.007) | .002 (-.004–.007) | .006 ( .002–.011) | .002 (-.004–.008) | **.003 (.001 – .006)** | *I^2^*= 0.0%  *p* = 0.50 |
| **SRI** |  |  |  |  |  |  |  |
| **Global Cognition** | Main effects | .051 (.021–.081) | .076 (.032–.120) | .010 (-.030–.051) | - | **.05 (.01-.08)** | *I^2^*= 58.0%  *p* = 0.09 |
|  | Interaction with follow-up time | -.002 (-.006–.002) | .006 (.001–.011) | .006 ( .001–.011) | - | .003 (-.002-.008) | *I^2^*= 71.9%  *p* = 0.03 |
| **IS** |  |  |  |  |  |  |  |
| **Global Cognition** | Main effects | .006 (-.025–.037) | .032 (-.013–.077) | -.038 (-.078–.002) | -0.013 (-.041–.015) | .00 (-.03-.02) | *I^2^*= 49.4%  *p* = 0.11 |
|  | Interaction with follow-up time | .001 (-.004–.005) | .006 (.000–.011) | .006 ( .001–.011) | .001 (-.005–.006) | **.003 (.000-.006)** | *I^2^*= 14.4%  *p* = 0.32 |

**Note:** All effect sizes are β[(95% CI] per 1-SD higher regularity per year. Models were adjusted for age, sex, race/ethnicity, study site, education, marital status, paid employment, body mass index, alcohol use, physical activity, smoking, vascular comorbidities, depressive symptoms, and use of sleep medication. Study-level estimates were pooled using random effects meta-analyses. Statistically significant pooled effect sizes (i..e. 95%CI does not include 0) are **bolded.** Cognitive tests are standardized and if applicable transformed to improve normality. Higher scores indicate better cognitive performance. RRI: Rest Regularity Index; SRI: Sleep Regularity Index; IS: Interdaily Stability MrOS: Osteoporotic Fractures in Men Study; RS: Rotterdam Study; MESA: Multi-Ethnic Study of Atherosclerosis; MAP: Rush Memory and Aging Project.
